# No benefits of prolonged vitamin D_3_ supplementation for adaptations to resistance training in old adults

**DOI:** 10.1101/2020.07.10.20150037

**Authors:** KS Mølmen, D Hammarström, K Pedersen, AC Lian Lie, RB Steile, H Nygaard, Y Khan, H Hamarsland, L Koll, M Hanestadhaugen, A Lie Eriksen, E Grindaker, JE Whist, D Buck, R Ahmad, TA Strand, BR Rønnestad, S Ellefsen

## Abstract

**Background:** Lifestyle therapy with resistance training is a potent measure to counteract age-related loss in muscle strength and mass. Unfortunately, many individuals fail to respond in the expected manner. This phenomenon is particularly common among older adults and those chronically diseased (e.g. chronic obstructive pulmonary disease, COPD), and may involve endocrine variables such as vitamin D. At present, the effects of vitamin D supplementation on responses to resistance training remain largely unexplored.

**Methods:** Ninety-five male and female participants (healthy, n=71; COPD, n=24; age 68 ± 5 years) were randomly assigned to receive either vitamin D_3_ or placebo supplementation for 28 weeks in a double-blinded manner (latitude 61°N, September-May). Seventy-eight participants completed the RCT, which was initiated by 12 weeks of supplementation-only (in average, 3333 IU.day^-1^), followed by 13 weeks of combined supplementation (2000 IU.day^-1^) and supervised whole-body resistance training (twice weekly), interspersed with testing and measurements. Outcome measures included multiple assessments of muscle strength (n=7), endurance performance (n=6), and muscle mass (n=3, legs, primary), as well as muscle quality (legs), muscle biology (*m. vastus lateralis*; muscle fibre characteristics, transcriptome), and health-related variables (e.g. visceral fat mass and blood lipid profile). For main outcome domains such as muscle strength and muscle mass, weighted combined factors were calculated from the range of singular assessments.

**Results:** Overall, 13 weeks of resistance training increased muscle strength (13% ± 8%), muscle mass (9% ± 8%) and endurance performance (one-legged, 23% ± 15%; whole-body, 8% ± 7%), assessed as weighted combined factors, and were associated with changes in health variables (e.g. visceral fat, − 6% ± 21%; [LDL]_serum_, −4% ± 14%) and muscle tissue characteristics such as fibre type proportions (e.g. IIX, −3%-points), myonuclei·fibre^-1^ (30% ± 65%), total RNA/rRNA abundances (15%/6-19%), and transcriptome profiles (e.g. ∼336 differentially expressed genes). Vitamin D_3_ supplementation led to robust increases in [25(OH)D]_serum_ (Δ49% vs placebo), but did not affect training-associated changes for any of the main outcome domains, with no interaction being evident with disease status or pre-RCT [25(OH)D]_serum_. In secondary analyses, vitamin D_3_ affected expression of gene sets involved in vascular functions in muscle tissue, strength gains in participants with high fat mass, and [cortisol]_serum_ (Δ20%), all of which advocate further study.

**Conclusions:** Vitamin D_3_ supplementation did not affect muscular responses to resistance training in old adults with or without COPD.

## Introduction

Aging is associated with progressive loss of muscle strength and mass, accompanied by declines in physical performance. In 2016, this had escalated into ∼11 million Europeans (> 65 years of age) suffering from sarcopenia,^1^ a formally recognized disease characterized by severe loss of muscle quantity and quality.^1^ Sarcopenia increases the likelihood of adverse events such as falling, fractures, physical disability, morbidity and mortality,^2,3^ further fuelling muscle deterioration, resulting in a spiralling decrease in overall health and health-related quality of life.^4–6^ In Europe, the prevalence of sarcopenia is expected to increase to at least ∼19 million by 2045,^1^ coinciding with increased proportions of older adults, potentiated by suboptimal nutrition and increasing incidences of causal morbidities such as systemic inflammatory diseases.^7,8^ For elderly to stay healthy, active and independent, efficient interventions are warranted for its prevention, treatment and reversal.^7,8^ To this end, lifestyle therapy with resistance training stand out as an attractive, low-cost and potent intervention.^9,10^ Unfortunately, the benefits of such interventions are not always consistent in the older population, with selected individuals and populations showing impaired abilities to increase muscle strength and mass.^11,12^ At present, this training-response-spectrum has an unknown causality, though it seems to interdepend on factors such as genetics,^13,14^ epigenetics,^14^ and composites of the inner physiological milieu, including nutrition,^15,16^ endocrine variables (e.g. vitamin D),^17,18^ and hallmarks of health such as low-grade chronic inflammation.^19^ This calls for development of combinatorial lifestyle protocols who, target and correct these factors alongside resistance training, allowing adequate muscle adaptations to occur.

Over the last two decades, vitamin D has emerged as a potential determinant of muscle functionality and biology.^20^ There seems to be a robust relationship between heterogeneity in vitamin D status and traits such as physical performance^(21–23)^ and susceptibility to falling,^24^ suggesting a causal association between vitamin D and increased risk of sarcopenia.^25^ As such, vitamin D status varies substantially in the human population, both in an annual cycle, and between individuals and groups of individuals.^26,27^ Vitamin D insufficiency is particularly prevalent in older adults, defined as 25-hydroxyvitamin D/25(OH)D) levels < 50 nmol. L^-1^, and especially in older adults living in the Northern Hemisphere,^27,28^ where cutaneous vitamin D synthesis is miniscule or absent during winter months.^29^ In accordance with this, exogenous vitamin D supplementation is gaining momentum as a potential ergogenic aid for preventing and treating sarcopenia.^25^ Despite this, its beneficial effects are ambiguous in existing intervention studies. While some studies and meta-analyses report favourable effects of vitamin D supplementation per se on muscle strength^30–32^ and falling,^33,34^ with benefits being more pronounced in subjects with low baseline values (< 30 nmol L^-1^)^35^ and in older subjects,^35^ others do not.^36–39^ These discrepancies may not be surprising, as resistance training is arguably necessary to provoke changes in muscle functions.^40^ However, a similar ambiguity is present in the few studies that have assessed the effects of vitamin D supplementation on outcomes of resistance training.^41–44^ While no study report clear benefits of vitamin D supplementation on alterations in muscle strength,^41–44^ muscle mass,^42–44^ or incidences of falling,^41,43^ a recent meta-analyses still concluded that it is beneficial for training-associated changes in lower body muscle strength.^40^

Consequently, we thus have limited and conflicting knowledge about the combined effects of vitamin D supplementation and resistance training on muscle functions and biology in humans. The present confusion may partly be attributed to methodological uncertainties in available studies, potentially lowering their ecological validity and explaining their lack of coherence with the resulting meta-analysis data. This includes heterogeneous study populations (varying from young adults^(42,44)^ to older adults^(44)^ to elderly^(41,43)^) with large differences in baseline 25(OH)D levels (average 31 nmol. L^-1 (43)^ – 71 nmol. L^-1 (44)^), large variation in vitamin D dosage (from 400 IU · day^-1 (43)^ – 4 000 IU · day^-1(42)^), lack of familiarization to strength tests^(41,43)^ (failing to ensure validity of data^(45)^), suboptimal training protocols^(41,43)^ (failing to comply with current recommendations of controlled maximal effort resistance training^(46,47)^), low compliance to training,^41,43^ and a lack of dietary assessment during the intervention.^41,43,44^ Also, neither of the studies included a period of vitamin D supplementation prior to resistance training, which may be necessary to prime muscle cells for adaptations, potentially acting by changing epigenetic traits, which has been observed in other cell types, such as T-cells^48^ and oral squamous cell carcinoma cells.^49^ Furthermore, the effects of vitamin D supplementation on muscle fibre characteristics and biology remain poorly understood and unclear.^50^ In theory, these effects can potentiate muscle fibres in two ways. Either directly by acting through vitamin D receptors in muscle fibres and progenitor cells, perhaps inducing intramuscular signalling pathways such as the p38 mitogen-activated protein kinase pathway,^51,52^ or indirectly by interacting with systemic signalling event, perhaps inducing testosterone signalling^(53)^ and thereby facilitating muscle plasticity. Our lack of insight is underlined by the longstanding uncertainty of the presence of vitamin D receptors in muscle tissue,^54^ though several indications suggesting its expression. First, there seems to be associations between mutations in the vitamin D receptor and muscle weakness in both humans and mice.^55,56^ Second, muscle-specific knock-out of the vitamin D receptor in mice deteriorates muscle strength and mass in a manner that resemble sarcopenia.^57^ The prevailing uncertainty is fuelled by a seeming lack of effects of vitamin D supplementation per se on the muscle transcriptome in vitamin D-insufficient frail elderly, though also in that study the vitamin D dosage was relatively low (400 IU. day^-1^).^58^ To date, a mere single study has assessed the effects of vitamin D supplementations on resistance-training induced muscle biological adaptations in humans, and as such assessing only a limited selection of traits and failing to disclose conclusive findings.^44^

The aim of the present study was to investigate the effects of 12 weeks of vitamin D_3_ supplementation only (3 333 IU. day^-1^), followed by 13 weeks of combined vitamin D_3_ supplementation (2 000 IU. day^-1^) and resistance training, on training-associated adaptations in a mixed population of older subjects. The study population includes individuals that were either at risk of developing sarcopenia (age or disease, i.e. COPD patients)^59,60^ or showed diagnostic indications of sarcopenia (16.4 % of the participants had appendicular skeletal muscle mass (kg)/m^2^ greater than two standard deviations below the sex-specific means of young adults).^61^ Outcome measures included a large range of muscle strength and endurance performance tests, multiple assessments of muscle mass, muscle quality, in-depth analyses of muscle biology including muscle fibre characteristics and analyses of the muscle transcriptome, and a range of health-related measures including body mass composition, blood variables and self-reported health variables.

## Methods

### Study ethics and participants

This study was approved by the Regional Committee for Medical and Health Research Ethics - South-East Norway (reference nr: 2013/1094) and was preregistered at clinicaltrials.gov (ClinicalTrials.gov Identifier: NCT02598830). All participants were informed about the potential risks and discomforts associated with the study and gave their informed consent prior to study enrolment. The study was conducted according to the *Declaration of Helsinki*.

Ninety-five male and female participants (age 68 ± 5 years, range 56-77) were included into the study (Figure 1). Eligibility criteria were consumption of less than 400 international units (IU) of vitamin D_3_ per day for the last two months leading up to the study, and either normal lung function or medical diagnosis of chronic obstructive lung disease (COPD; GOLD^(62)^ grade II or III, FEV_1_ predicted between 80 % − 30 %, FEV1/FVC < 70 % after reversibility testing with inhalation of salbutamol and ipratropiumbromid). Exclusion criteria were unstable cardiovascular disease, chronic granulomatous disease, known active malign illness the last five years, serious psychiatric comorbidity, steroid use the previous two months and musculoskeletal disorders preventing the participant to follow the resistance training program. Initially, all participants were screened using spirometry and a medical questionnaire. Healthy subjects were included based on these data. A medical doctor consulted participants with spirometry values corresponding to GOLD grade II or III prior to inclusion. All participants were recreationally active but had not partaken in systematic resistance training for the previous 12 months. During the study conduct, all subjects were instructed to minimize vitamin D intake to < 400 IU. day^-1^ and to abstain from solarium and travels to southern and/or sunny areas.

**Figure 1.**
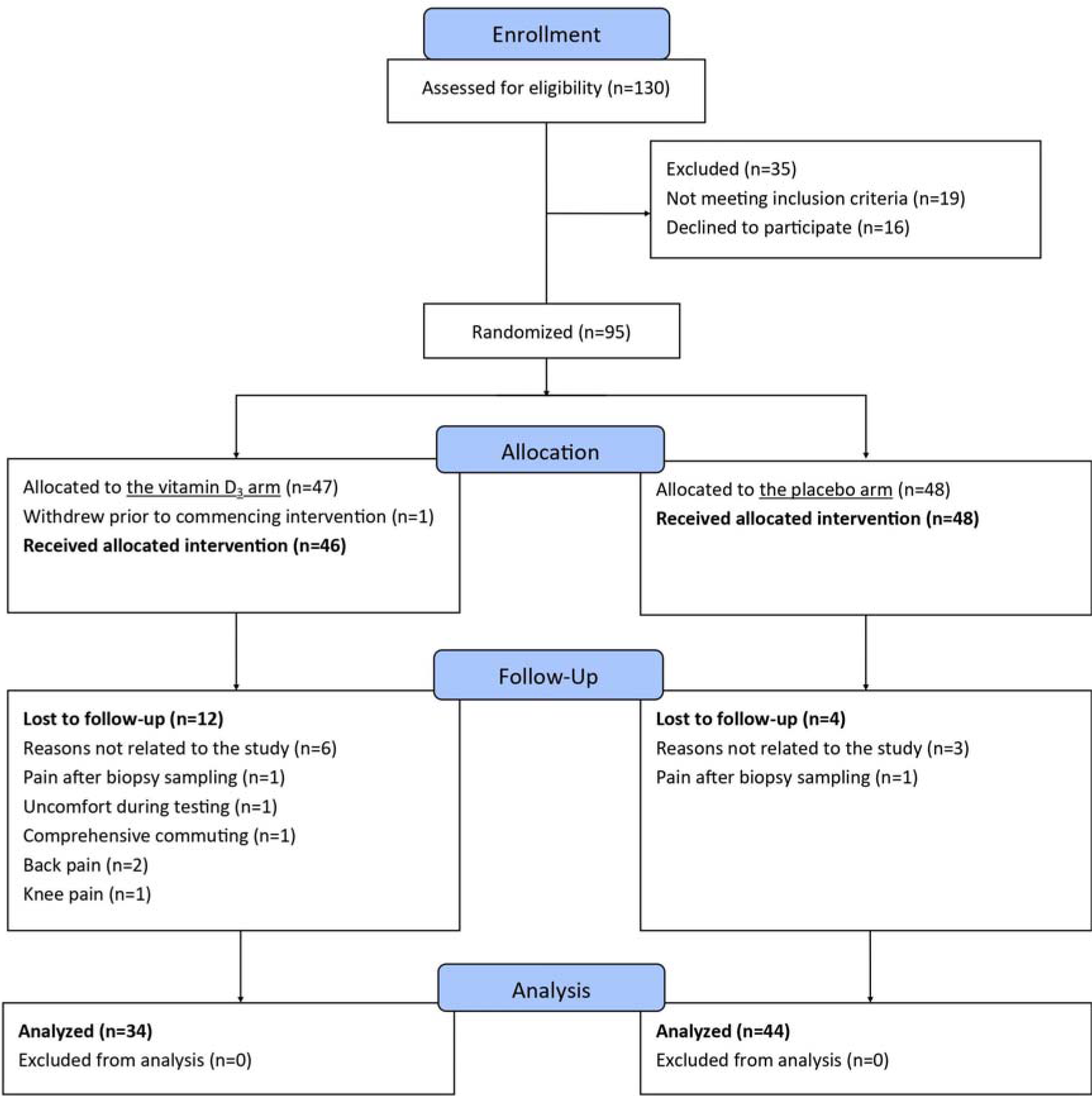
CONSORT flow chart of the study.

Participants were randomly assigned into one of the two study arms (vitamin D_3_ vs placebo) using concealed allocation, stratified by sex and health status (COPD vs non-COPD) (Figure 1 and Table 1). An off-site third party performed the randomization. During the initial two weeks of the study, the vitamin D_3_ arm consumed 10 000 IU vitamin D_3_. day^-1^, followed by 2 000 IU. day^-1^ for the rest of the study period. Placebo capsules contained cold-pressed olive oil and were identical in appearance to vitamin D_3_ capsules. Pharma Nord ApS (Vejle, Denmark) produced the two supplements, complying with Good Manufacturing Practice requirements. All participants consumed 500 mg calcium. day^-1^ (Nycoplus, Takeda AS, Asker, Norway). Vitamin D status was assessed as 25(OH)D levels in blood (Figure 2), corroborating with previous studies.^63^ 25(OH)D levels are not influenced by parathyroid hormone (PTH) and is more stable than the metabolically active form of vitamin D; 1,25 dihydroxycholecalciferol.^64^

**Table 1.**
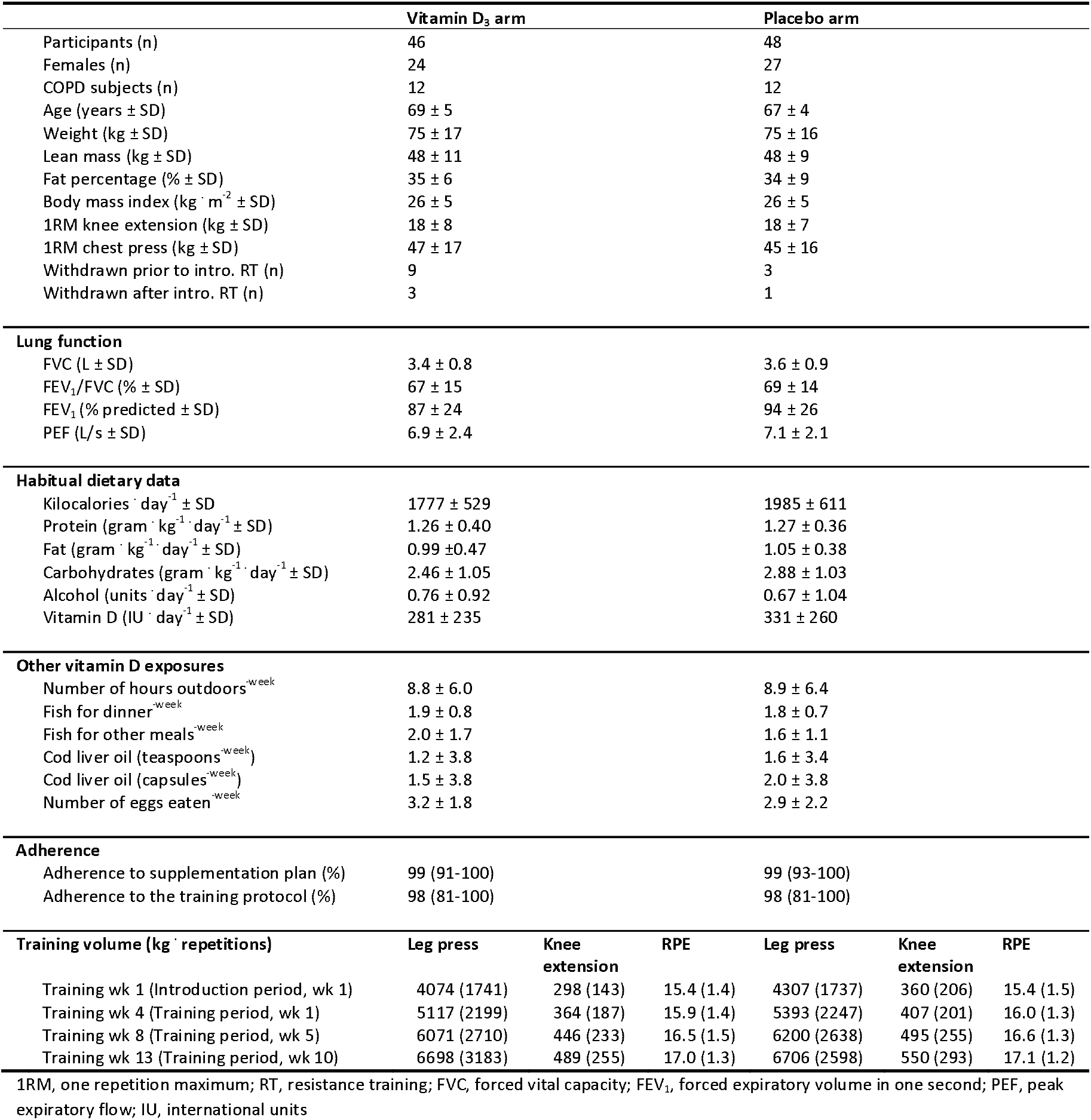
Participant characteristics.

**Figure 2.**
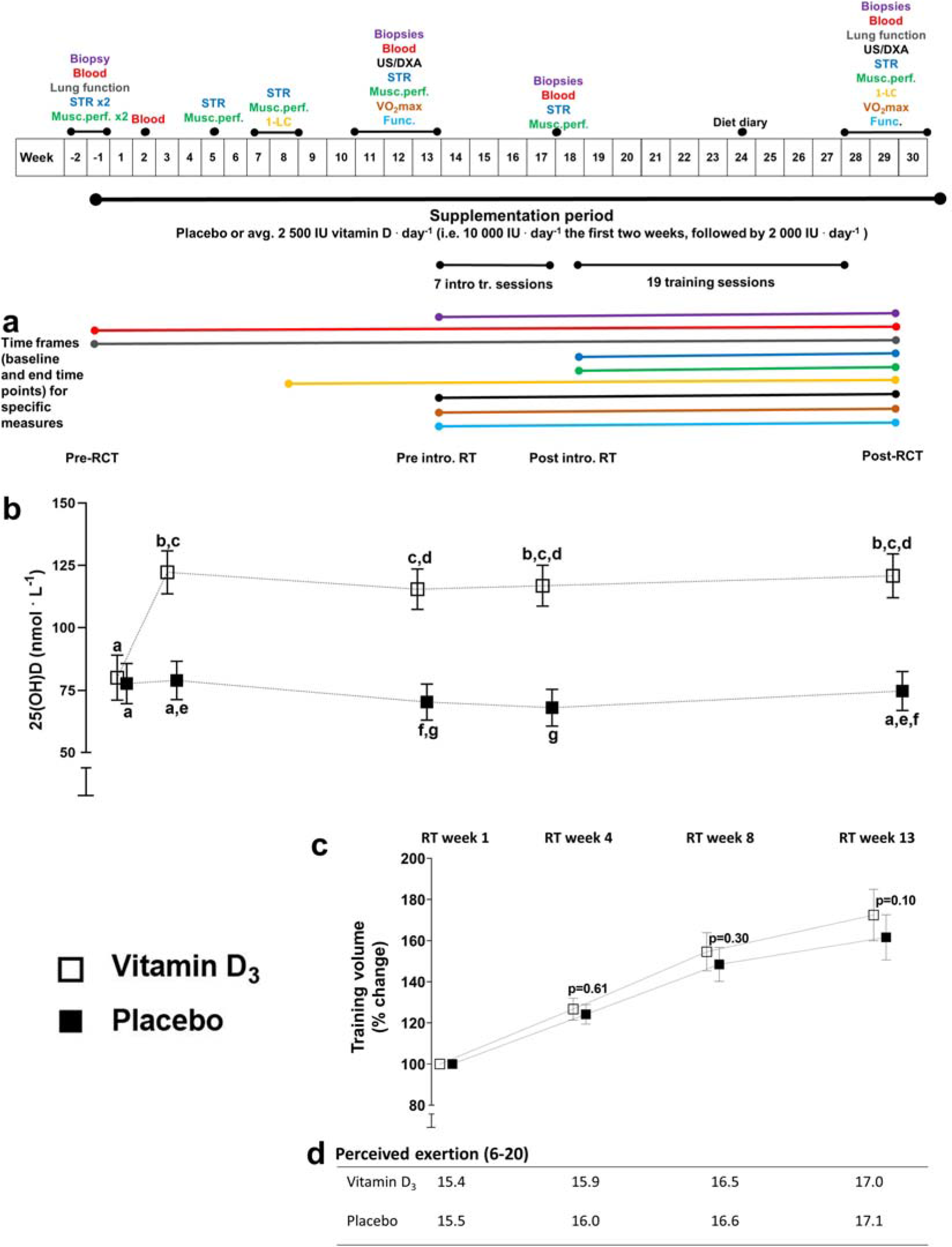
Schematic overview of the study protocol. Pre-defined main time frames (baseline and end time points) for specific outcome measures (the colour lines represents the measurement marked with the same colour at the top of the figure; a), vitamin D-status during the RCT (b), training volume during the resistance training intervention (c), and perceived exertion (Borg RPE, 6-20) reported after training sessions (d). The training volume was calculated as average increase in volume (kg. repetitions) in leg press and knee extension from the first week of training. STR, maximal strength test; Musc.perf., test of muscular performance; 1-LC, one-legged cycling test; Func., test of functional capacity (6-minute step test and 1-minute sit-to-stand test); US, ultrasound measures of muscle thickness; DXA, Dual-energy X-ray Absorptiometry; VO_2_max, maximal oxygen uptake; IU, international units; RT, resistance training; 25(OH)D, calcifediol. In (b), statistical differences between time points and supplementation arms are denoted by letters: different letter indicates p < 0.05, i.e. all time point measures denoted with the same letter are statistically similar (p > 0.05). Data for 25(OH)D and training volume are presented as means with 95 % confidence intervals.

Of the 95 participants included in the study, 1 withdrew from the study prior to onset on supplementation, 12 withdrew prior to onset of resistance training (vitamin D_3_ arm, n = 9; placebo arm, n = 3), and 4 participants withdrew during the resistance training period (vitamin D_3_ arm, n = 3; placebo arm, n = 1) (Figure 1). In summary, 78 participants completed the study; 58 healthy participants and 20 COPD participants. For participant characteristics, see Table 1.

Insert Figure 1 and Table 1 around here

### Study conduct

The study was conducted as a double-blind randomized clinical trial (RCT), consisting of an initial 12 weeks of supplementation-only (in average, 3 333 IU vitamin D_3_. day^-1^ or placebo), followed by 13 weeks of combined supplementation (2 000 IU vitamin D_3_. day^-1^ or placebo) and resistance training (Figure 2). Neither the participants nor the investigators did know which supplement the participants were randomized to. Unblinding was performed after completion of analyses of the primary outcome measures. The intervention was conducted at Lillehammer, Norway (latitude 61° N) from September to May, ensuring low or no natural vitamin D synthesis by the skin from sunlight UVB radiation during the study period.^29^ Prior to onset of the supplementation protocol (i.e. pre-RCT), participants undertook a two-week period of baseline testing and tissue/blood sampling (Figure 2, Weeks −2 and −1), including testing of lung function, unilateral strength and muscle performance (duplicated testing for unilateral strength and muscle performance, separated by at least 48 hours; the highest values achieved during the second test was used in further analyses, as the first test was conducted at ∼95 % of maximal effort), and collection of fasting blood and rested-state muscle biopsy, sampled from m. vastus lateralis of the dominant leg using the microbiopsy procedure (Bard Magnum, Bard, Covington, GA, USA). Thereafter, participants were randomized to the two supplementation arms. After two weeks of supplementation, a second blood sample was collected (Figure 2, Week 2) to validate the efficacy of vitamin D_3_ supplementation on blood 25(OH)D levels. Prior to introduction to resistance training, the participants conducted repeated performance tests at several occasions (Figure 2, Week −2 - Week 13), including unilateral maximal strength and muscular performance, isokinetic unilateral knee-extension torque, measures of functional capacity (i.e. 6-minute step and 1-minute sit-to-stand test), submaximal and maximal one-legged cycling, and maximal bicycling. In the week prior to introduction to resistance training (Figure 2, Week 13), we also collected bilateral rested-state biopsies and a fasted blood sample, measured muscle thickness of *m. vastus lateralis* and *m. rectus femoris* using ultrasound (SmartUs EXT-1M; Telemed, Vilnius, Lithuania), and assessed body-mass composition using dual-energy X-ray absorptiometry scan (DXA; Lunar Prodigy, GE Healthcare, Chicago, IL, USA).

The training intervention consisted of 13 weeks of two weekly full-body resistance training sessions (Figure 2, Week 14-27). Leg exercises were performed unilaterally to allow within-participant differentiation of resistance training load. Accordingly, for each participant, the two legs were randomly assigned to perform either three sets with 10 repetitions to exhaustion (high-load resistance exercise) or three sets with 30 repetitions to exhaustion (low-load resistance exercise); i.e. each participant performed both protocols in each session. For the upper-body, resistance exercises were performed bilaterally, consisting of two sets of 10 repetitions to exhaustion. After seven training sessions (i.e. after 3.5 weeks of training; post-introduction to resistance training), participants performed a selected battery of tests and measurements (Figure 2, i.e. Week 17-18), including rested-state bilateral muscle biopsies, a fasted blood sample, and measures of muscle strength, performance and torque. This enabled assessment of the initial responses to resistance training and ruled out the initial neural adaptations to resistance training for performance tests. After the training intervention (i.e. post-RCT), the complete battery of tests and measurements were repeated (Figure 2, i.e. Week 28-30). In week 24, participants conducted a dietary registration, in which they logged their dietary intake for three days, including one weekend day (Table 1). Throughout the entirety of the study, participants completed a weekly health survey, including information about experienced health and potential discomforts with nutritional supplementation, i.e. digestion problems, sleep problems, issues with the urinary system, issues with the vestibular system and dermal irritations. Moderate verbal motivation was given to all participants during all performance tests.

### Resistance-exercise training protocol

All participants performed the same whole-body resistance-exercise training program, consisting of the following exercises (named in the order of conductance): unilateral leg press, unilateral knee extension, unilateral knee flexion, chest press and lat pulldown. Leg exercises were performed as three series of 10 repetitions (high-load) and 30 repetitions (low-load) to exhaustion (10RM and 30RM, respectively), and upper-body exercises were performed as two series of 10 repetitions (high-load) to exhaustion, as previously described. Exercises and sets were separated by two minutes of rest. For leg exercises, all three sets were concluded on one leg before the other leg was exercised. The exercise order of the leg performing high-load and low-load training was changed for each session. For all exercises, training loads were adjusted from session to session, i.e. when participants managed to perform more than 12 or 35 repetitions per set for high- and low-load training, respectively. All sessions were supervised by qualified personnel to ensure correct technical execution and to ensure maximal efforts through verbal encouragement. To aid recovery and to ensure adequate protein intake after training, participants ingested half a protein bar immediately after each training session (∼15g protein; Big 100, Proteinfabrikken, Sandefjord, Norway).

### Spirometry

Spirometry testing was performed using either the Oxycon Pro™ with the TripleV digital volume sensor (Carefusion GmbH, Höchberg, Germany) or the Spirare SPS320 ultrasonic spirometer (Diagnostica AS, Oslo, Norway) following guidelines from the American Thoracic Society and the European Respiratory Society.^65^ Importantly, for each particular participant, all spirometry tests were performed using the same system. Participants with COPD were tested before and after inhalation of two bronchodilators (salbutamol, 0.2 mg and ipratropiumbromid, 20 µg).

### Muscle strength and performance

Maximal muscle strength was assessed as one repetition maximum (1RM) in unilateral knee extension and leg press (Technogym, Cesena, Italy) and bilateral chest press (Panatta, Apiro, Italy). Each test started with specific warm-up, consisting of 10, 6 and 3 repetitions at 40, 70 and 85 % of the anticipated maximum. Thereafter, 1RM was found by increasing the resistance progressively until the weight could not be lifted through the full range of motion. Loads were increased in intervals of 1.25, 2.5 and 1.25 kg for knee extension, leg press and chest press, respectively. Two minutes of rest was provided between attempts. Maximal handgrip strength was measured for the dominant hand using a hand-held dynamometer (Baseline®, Fabrication Enterprises, Inc., Elmsford, NY, USA). Each test session consisted of three attempts, and the average score was used in further analyses.

Muscle performance was defined as the number of repetitions achieved at 50 % of pre-RCT 1RM and was assessed in unilateral knee extension and bilateral chest press. Participants were instructed to lift at a composed and controlled pace, with < 1 second breaks in the lower and upper position. When this requirement could not be met, or participants failed to lift the weight through the full range of motion, the test was aborted.

Isokinetic unilateral knee-extension torque was assessed using a dynamometer (Humac Norm, CSMi, Stoughton, MA, USA). Participants were seated and secured with the knee joint aligned with the rotation axis of the dynamometer. Maximal isokinetic torque was tested at three angular speeds (60°, 120° and 240°. s^-1^) with two minutes of rest provided between each of them. Prior to each test session, participants were familiarized with the test protocol by performing three submaximal efforts at each angular speed. Participants were given three attempts performed in immediate succession. The highest value was used in further analyses.

For all tests of unilateral strength and performance, the dominant leg was tested first. Seat position and general settings for each test were noted for each participant and reproduced at each time-point.

### One-legged cycling and bicycling performance

Participants conducted one-legged cycling tests (Excalibur Sport, Lode BV, Groningen, the Netherlands) to assess O_2_-costs of submaximal cycling, and maximal one-legged oxygen consumption (VO_2_max) and power output (Wmax). Each test was initiated by two x 5 min submaximal workloads at 30 and 40 watts (healthy), respectively, or 20 and 30 watts (COPD) with a cadence of 60 revolutions per minute (rpm). Loads were individually adjusted if the predefined workload was higher than 50 % of the Wmax achieved during the familiarization session. Thereafter, a maximal step-wise incremental protocol was conducted (10 and 5 watts. min^-1^ for healthy and participants with COPD, respectively). Starting loads were individually adjusted to elicit exhaustion after 6-10 min of cycling, based on results from the familiarization session. The cadence was freely chosen (> 50 rpm). The test was terminated when cadence fell below 50 rpm. For all participants, submaximal and maximal performance on the dominant leg was tested first. After testing of the first leg, participants were allowed 20 minutes rest and/or low-intensity cycling, before testing of the other leg. During one-legged cycling tests, a 10 kg counterweight was attached to the contralateral ergometer crank to facilitate smooth cycling. The foot of the non-exercising leg was rested on a chair placed in front of the subject. Breath-to-breath measurements of pulmonary oxygen consumption and ventilation (JAEGER Oxycon PRO™; Carefusion GmbH, Höchberg, Germany) and heart rate (Polar Electro Oy, Kempele, Finland) was monitored continuously during all tests. The average oxygen consumption during the last two minutes of each submaximal workload was defined as the O_2_-cost, while VO_2_max was defined as the highest average oxygen consumption measured over a period of 30-s. Measurement of capillary lactate concentration (Biosen C-line, EKF Diagnostics, Barleben, Germany) was performed after finalization of tests.

Testing of maximal bilateral cycling VO_2_max and Wmax was performed on a separate day. A step-wise incremental protocol (20 and 15 watts. min^-1^ for healthy men and women, respectively; 10 watts. min^-1^ for participants with COPD) was conducted. Oxygen consumption was measured continuously using a computerized metabolic system with mixing chamber (JAEGER Oxycon PRO™; Carefusion GmbH, Höchberg, Germany). Prior to each cycling test, the gas analyser was calibrated using certified calibration gases with known concentrations, and the flow turbine (TripleV; JAEGER, Carefusion GmbH, Höchberg, Germany) was calibrated using the metabolic system’s automatic volume calibration, or a 3 L, 5530 series calibration syringe (Hans Rudolph Inc., Kansas City, MO, USA), for one-legged and bicycling tests, respectively.

### Functional performance

One-min sit-to-stand and 6-min step tests were conducted in consecutive order on the same test day. Each test session was initiated with 10 min warm-up of low-intensity bicycling. Briefly, during the 1-minute sit-to-stand tests, participants were instructed to fold their arms and sit/stand up for as many times possible during a 1-min period. The seat was 0.45 m from the floor. Sit-to-stand repetitions were approved if both knees and hip joints were fully extended after each seating. Three minutes after the 1-minute sit-to-stand test, the 6-min step test was conducted. Briefly, participants were instructed to perform as many steps as possible onto a 20 cm high step box with a non-slip rubber surface within six minutes (Reebok Step; Boston, MA, USA). During each step, participants were instructed to place both legs on the box, with the hip fully extended.

### Muscle thickness by ultrasound and dual-energy X-ray absorptiometry-derived body mass measures

Prior to measurements of muscle thickness and DXA measurements, the participants were instructed to attend an overnight fast and avoid heavy physical activity for the last 24 h leading up to the event.

Muscle thickness of *m. vastus lateralis* and *m. rectus femoris* were measured using B-mode ultrasonography (SmartUs EXT-1M, Telemed, Vilnius, Lithuania) with a 39 mm 12 MHz, linear array probe. Transverse images were obtained ∼60 % distally from the *trochanter major* towards the femoral lateral epicondyle. Three images were captured for each muscle, where the probe was relocated to the same position between each image. The position of the probe was marked on the skin and subsequently marked on a soft transparent plastic sheet superimposed on the thigh. Landmarks such as moles and scars were also marked on the plastic sheet for relocation of the scanned areas during post-training measurements. During analysis, pre and post images from the same participant were analyzed consecutively using the Fiji software,^66^ and by two independent researchers. The average muscle thickness of the three images captured per muscle was used for further analyses.

Body composition was determined using DXA (Lunar Prodigy, GE Healthcare, Madison, WI, USA) and was analyzed using the manufacturer’s software, in accordance with the manufacturer’s protocol. Leg lean mass was defined as the region distally of *collum femoris*. Care was taken to match the region of interest on pre and post images. Analyses of both muscle thickness and body composition were performed in a blinded manner regarding participant identity and time point of the measurement.

### Blood sampling and measurements, and muscle biopsy sampling

Prior to collection of blood and muscle biopsies, participants were instructed to attend an overnight fast and to avoid heavy physical activity for the last 48 h leading up to the event. Blood samples were collected from an antecubital vein into serum-separating tubes and kept at room temperature for 30 min before centrifugation (2600 g, 15 min). Serum was aliquoted and stored at −80°C until further processing. Serum concentrations of total testosterone, cortisol, growth hormone, insulin-like growth-factor 1 (IGF-1), sex-hormone binding globulin (SHBG) and androstenedione were measured using an Immulite 2000 analyzer with kits from the Immulite Immunoassay System menu (Siemens Medical Solutions Diagnostics, Malvern, PA, USA). Serum 25(OH)D, parathyroid hormone, calcium, albumin, creatinine, creatine kinase, aspartate aminotransferase, C-reactive protein, triglycerides, low-density lipoprotein, high-density lipoprotein, thyroid hormones and iron metabolism variables were measured using a Roche Cobas 6000 analyzer and kits from Roche (Roche Diagnostics, Rotkreuz, Switzerland).

Muscle biopsies were obtained from *m. vastus lateralis* under local anaesthesia (Lidocaine, 10 mg. ml^-1^, AstraZenaca AS, Oslo, Norway) using a 12-gauge needle (Universal Plus, Medax, San Possidonio, Italy) operated with a spring-loaded biopsy instrument (Bard Magnum, Bard, Covington, GA, USA), as previously described.^67^ The biopsies were sampled at 1/3 of the distance from the patella to the *anterior superior iliac spine*. The tissue was quickly dissected free of blood and visible connective tissue in ice-cold sterile saline solution (0.9% NaCl). Samples for immunohistochemistry were transferred to a 4 % formalin solution for fixation for 24-72 h, before further preparation. Samples to RNA analyses were blotted dry, snap-frozen in isopentane (−80°C) and stored at −80°C until further prosessing.

### Immunohistochemistry

Formalin-fixed muscle biopsies were processed rapidly using a Shandon Excelsior ES (Thermo Fisher Scientific, Waltham, MA, USA), whereupon biopsies were paraffin-embedded and sectioned into transverse sections (4 µm). Antigen retrieval was performed at 97°C for 20 min in a target retrieval solution (cat.no. DM828, Agilent Dako, Santa Clara, CA, USA) using a PT link (PT 200, Agilent Dako, Santa Clara, CA, USA). Staining was performed using a DAKO Autostainer Link 48 (Agilent Dako, Santa Clara, CA, USA). For determination of muscle fibre types, cross-sections were first treated with protease 2 (cat.no. 760-2019, Roche Diagnostics, Rotkreuz, Switzerland), before they were triple-stained using 2.5 µg. ml^-1^ BA-F8, BF-35 and 6H1 (all from Developmental Studies Hybridoma Bank, University of Iowa, Iowa City, IA, USA; BA-F8 and BF-35 deposited by Schiaffino, S., Uni. of Padova, Italy; 6H1 deposited by Lucas, C., Uni. of Sydney, Australia). Visualization of the primary antibodies was achieved by incubation of appropriate secondary antibodies, diluted 1:400: goat anti-mouse Alexa Fluor (Thermo Fisher Scientific, Waltham,MA, USA) 350 (IgGγ2b, cat.no. A21140), 488 (IgGγ1, cat.no. A21121) and 594 (IgM H+L, cat.no. A21044) for BA-F8, BF-35 and 6H1, respectively.

For determination of muscle fibre cross-sectional area (CSA) and the number of myonuclei per muscle fibre type, a different tissue cross-section was double stained using primary antibodies against muscle fibre membrane (dystrophin, diluted 1:100, cat.no. PA1-21011; Thermo Fisher Scientific, Waltham, MA, USA) and myosin heavy chain I (diluted 1:2000, cat.no. M8421, Sigma-Aldrich, Saint-Louis, MO, USA). Visualization was achieved using the secondary antibodies Alexa Fluor 594 (IgG H+L, diluted 1:400, cat.no. A11037) and 488 (IgG1γ1, diluted 1:400, cat.no. A21121), respectively (Thermo Fisher Scientific, Waltham, MA, USA). Muscle sections were then covered with a coverslip and glued with EverBrite™ Hardset Mounting Medium with DAPI (cat.no. 23004, Biotium Inc., Fremont, CA, USA), to visualize cell nuclei.

Images of stained cross-sections were captured using a high-resolution camera (Axiocam, Zeiss, Oberkochen, Germany) mounted on a light microscope (Axioskop-2, Zeiss, Oberkochen, Germany), with a fluorescent light source (X-Cite 120, EXFO Photonic Solutions Inc., Mississauga, Canada). Multiple images were taken using 20x objectives to capture the entirety of each cross-section. For representative images, see Figure 3. All analyses of muscle fibre characteristics were performed using automated procedures, ensuring unbiased quantification.

**Figure 3.**
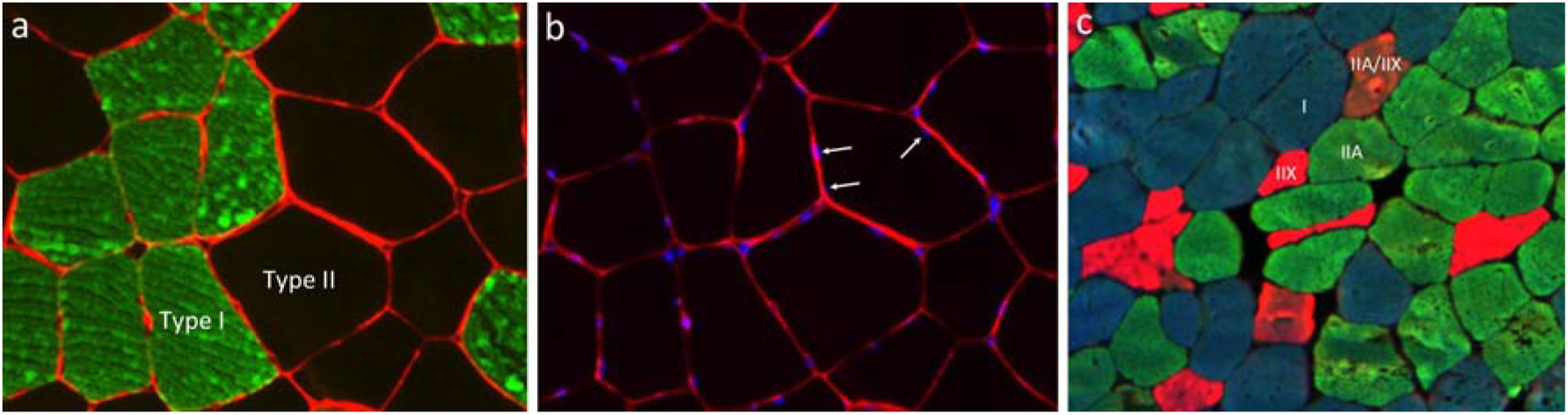
Representative immunohistochemistry images of (a) myosin heavy chain I (green) and cell membrane (red), (b) myonuclei (blue) and cell membrane (dystrophin, red), and (c) myosin heavy chain I (blue), IIA (green), IIX (red), and IIA/IIX hybrids (orange). Images in (a) and (b) are from the same tissue cross-section: triple-staining myosin heavy chain I, dystrophin and cell nuclei.

Analyses of muscle fibre type proportions were performed using the Cell Counter function in the Fiji software,^66^ whereby muscle fibres were categorized as either type I, type IIA, type IIX or hybrid fibres type IIA/IIX. Sections and/or images with insufficient staining to distinguish between fibre types were excluded. Muscle fibre type-specific CSA (type I or type II) were calculated using the TEMA software (CheckVision, Hadsund, Denmark). Myonuclei were counted using the CellProfiler software.^68^

### Total RNA extraction and qPCR

Approximately 10-20 mg of wet muscle tissue (average 13 ± 4 mg, range 3 – 26 mg) was homogenized in a total volume of 1 ml TRIzol reagent (Invitrogen, Carlsbad, CA, USA) using 0.5 mm RNase-free zirconium oxide beads and a bead homogenizer (Bullet Blender, Next Advance, Averill Park, NY, USA), as previously described.^67^ To enable analysis of target gene expression per unit tissue weight, an exogenous RNA control (λ polyA External Standard Kit, Takara Bio Inc., Shiga, Japan) was added at a fixed amount (0.04 ng. ml^-1^ of Trizol reagent) per extraction prior to homogenization, as previously described.^69,70^ Following phase separation, 450 µl of the upper phase was transferred to a new tube and RNA was precipitated using isopropanol. The resulting RNA pellet was washed three times with 75 % ethanol and eluted in TE buffer to 100 ng RNA. µl^-1^, following quantification of total RNA concentration using µDrop plate and the Multiskan GO microplate spectrophotometer (Thermo Fisher Scientific, Waltham, MA, USA). RNA integrity was assessed using capillary electrophoresis (Experion Automated Electrophoresis Station using RNA StdSens Assay, Bio-Rad, Hercules, CA, USA) with average integrity score (RNA quality indicator; RQI): 8.9 ± 0.8.

Five hundred nanograms of RNA were reverse transcribed using anchored oligo-dT (Thermo Fisher Scientific, Waltham, MA, USA), random hexamer primers (Thermo Fisher Scientific, Waltham, MA, USA) and Super-Script IV Reverse Transcriptase (Invitrogen, Carlsbad, CA, USA) according to manufacturers’ instructions. All samples were reverse transcribed in duplicates and diluted 1:50 prior to quantitative real-time polymerase chain reaction (qPCR). qPCR reactions were conducted using a fast-cycling real-time detection system (Applied Biosystems 7500 fast Real-Time PCR Systems, Life Technologies AS), with total volumes of 10 μl, containing 2 μl cDNA (1:25 dilutions), target gene-specific primers (final concentration 0.5 μM) and a commercial master mix (2X SYBR Select Master Mix, Applied Biosystems, Life Technologies Corp., Carlsbad, CA, USA). qPCR reactions consisted of 40 cycles (3 s 95°C denaturing and 30 s 60°C annealing). Melt-curve analyses were performed for all reactions to verify single-product amplification. Gene-specific primers were designed using Primer3Plus^71^ and synthesized by Thermo Scientific, except for the external RNA control, for which primers were supplied with the kit (λ polyA External Standard Kit, Takara Bio Inc., Shiga, Japan). Raw fluorescence data were exported from the platform-specific software and amplification curves were modelled using a best-fit sigmoidal model using the qpcR-package^72^ written for R.^73^ Threshold cycles (Ct) were estimated from the models by the second-derivate maximum method with technical duplicates modelled independently. Amplification efficiencies were estimated for every reaction.^74^ For every primer pair, mean amplification efficiencies (*E*) were utilized to transform data to the linear scale using *E*^−Ct^. Primer sequences and primer characteristics (i.e. average primer efficiencies and Ct values) are presented in Table S1. Gene expression data were log-transformed prior to statistical analysis. As Ct values, but not primer efficiencies depend on RNA integrity,^75^ RQI scores were used as a random variable on a *per*-target basis to control for potential degradation during statistical analyses (see below).

### RNA sequencing

RNA sequencing was performed on pairwise muscle samples collected before the RCT (vitamin D_3_, n = 11; placebo, n = 13), after 12 weeks of supplementation-only (vitamin D_3_, n = 24; placebo, n = 29), after 3.5 weeks of introduction to resistance training (vitamin D_3_, n = 23; placebo, n = 28), and after 13 weeks of resistance training (vitamin D_3_ arm, n = 24; placebo arm, n = 29). Samples was selected based on quality of total RNA samples (RQI > 7.0, avg 9.0 ± 0.5). Participants with complete sets of muscle biopsies were prioritized. For each muscle sample, mRNA sequencing libraries were prepared from 1000 ng of total RNA using TruSeq Stranded Total RNA Library Prep (Illumina, San Diego, CA, USA). Paired-end sequencing (150 bp) was performed using an Illumina HiSeq 3000 (Illumina, San Diego, CA, USA) at the Norwegian Sequencing Centre, Oslo, Norway.

### Data analyses and statistics

As defined in the pre-registration of the study protocol (ClinicalTrials.gov Identifier: NCT02598830), investigation of the effects of vitamin D_3_ supplementation on the diverse set of research questions and outcome measures was performed using different defined baseline time points (outlined in Figure 2). For transparency, statistical comparisons of all outcome measures and all relevant time points are presented in Table S2 and S3. These tables also specify the statistical models used for each specific variable and analysis. In general, for continuous variables, the effects of vitamin D_3_ supplementation (compared to placebo) were investigated using linear mixed-effects models with the relative change from baseline being defined as the dependent variable and the supplementation arms being defined as the fixed effect. The two different training loads (high- and low-load) were added to the model as repeated measures/observations (for unilateral outcome measures), and baseline values were used as co-variates. For all participants, random intercepts were specified. For all unilateral leg variables, interaction effects were explored between the fixed effect and health status (COPD vs. non-COPD) and training loads. For other variables, interactions were investigated between the fixed effect (vitamin D_3_ vs placebo) and health status, with the exception for blood variables, for which the interaction with sex was also examined. For all statistical analyses of immunohistochemical variables (muscle fibre CSA, fibre type proportion, and myonuclei. fibre^-1^), the models were weighted for the number of counted fibres per biopsy. This was done to account for the reduced reliability accompanying fewer observations/fibres (see Figure S2). For non-continuous variables, a different statistical approach was used to investigate the effects of the vitamin D_3_ supplementation. For fibre type proportions (immunohistochemistry) and variables from the weekly health diary, a generalized linear mixed model (GLMM) with binomial error distribution and link function was used to examine differences in changes between supplementation arms (time * supplementation arm interactions). For gene family-based analyses of myosin heavy-chain mRNA data,^76^ a GLMM with negative binomial distribution/link function (log-link) was used following transformation to transcript counts.^77^ Target gene mRNA abundance, expressed as per unit muscle weight using the external reference gene, were analyzed using mixed linear models with within-model normalization through the addition of random effects of technical replicates. To allow for gene-specific variances, variance functions were specified per strata (per gene). RQI scores were included in the model on a per target basis to control for RNA degradation. The number of observations per statistical analysis is presented in Table S2. For most outcome measures, the main effect of time was examined using mixed models, using absolute values for the dependent variable and time points as repeated measures/observations with random intercepts for each subject (Table S2 for complete overview).

### Transcriptome analyses

Gene counts were modelled using negative binomial GLMM with the total library size modelled as a fixed effect together with study conditions (time point and supplementation arms).^78^ In addition, a model containing the amount of tissue used in library preparations as an offset term was used to account for fluctuations in RNA-to-tissue ratios, as fixed amounts of total RNA was used to normalize samples during cDNA synthesis. The effect of resistance training on gene counts was assessed as i) the effect of time and ii) the effect of supplementation arm (vitamin D_3_ and placebo supplementation). For analyses of the effect of time, differential expression was evaluated using GLMMs containing only the time factor, combining all data irrespective of supplementation arm. For analyses of the effect of supplementation over time, differential expression was evaluated using GLMMs containing the interaction between time and supplementation arm. In all models, a single random effect was used, giving each participant an individual intercept. Models were iteratively fitted using glmmTMB.^79^ Genes were identified as differentially expressed when the absolute log_2_ fold-change was greater than 0.5 and the adjusted P-value (false discovery rate adjusted per model coefficient) was below 5 %. Enrichment analyses of gene ontology (GO) gene sets were performed using two approaches. First, a non-parametric rank test^(80,81)^ was performed based on gene-specific minimum significant differences (MSD). MSD was defined as the lower limit of the 95 % confidence interval (CI, based on estimated standard errors) around the log fold-change (FC) when log(FC) > 0 and the negative inverse of the upper 95 % CI when log(FC) < 0. Genes with MSD < 0 were further ranked based on *P*-values. The rank test assessed non-directional changes in gene sets. Second, gene set enrichment analysis (GSEA)^82^ was performed to quantify directional regulation of the gene set. GSEA was performed using the fgsea package,^83^ with - log_10_(p-values) * log_2_(fold-change) acting as the gene level metric.^84^ Consensus results were given higher importance. GO gene sets (biological process, cellular component and molecular function) were retrieved from the molecular signature database (version 7.1).^85^ Overview of GO analyses with exact p-values are presented in Table S5, S6 and S7.

To achieve reliable assessment of main outcome domains, and thus to lower the risk of statistical errors, combined factors were calculated for outcome measures relating to muscle strength, muscle mass, one-legged endurance performance and whole-body endurance performance, respectively. For complete overview over the composition of each factor, see Table S4. During factor calculation, each of the underlying variables were normalized to the participant with the highest value recorded during the RCT, resulting in individual scores ≤ 1. Thereafter, outcome domain factors were calculated as the mean of the normalized values for each variable for each subject (e.g. the muscle mass factor of the legs included muscle thickness, lean leg mass, and muscle fibre CSA). To evaluate the biological coherence of these factors, a factor analysis was performed to ensure correlation between the combined factors and their underlying outcome variables (Table S4).^86^ To assess the effect of vitamin D_3_ supplementation on changes of these combined factors, linear mixed-effects models were used, as previously described. In addition, these factors were used to investigate the influence of pre-RCT levels of 25(OH)D, body fat proportions and body mass index on the effects of vitamin D_3_ supplementation. To perform these analyses, each of the two supplementation arms were divided into quartiles, defined by baseline 25(OH)D, body fat percentage and body mass index levels, respectively (quartile 1, lowest,…quartile 4, highest). For each of the calculated factors, the effect of quartile and the interaction between quartile and supplementation arm was examined using mixed modelling.

Statistical significance was set to p < 0.05. In the text, data are presented as means ± standard deviation. In figures, data are shown as adjusted, estimated marginal means of relative changes and differences in relative changes between supplementation arms, with 95 % confidence intervals, unless otherwise stated. Statistical analyses were performed using SPSS Statistics package version 24 (IBM, Chicago, IL, USA) and R software.^73^ Figures were made using Prism Software (GraphPad 8, San Diego, CA, USA) and R software.^73^

## Results and discussion

### Effects of vitamin D_3_ supplementation on 25(OH)D in blood

At pre-RCT, participants in vitamin D_3_ and placebo intervention arms had similar [25(OH)D] levels in serum (80 nmol · L^-1^ vs 78 nmol · L^-1^, range: 24-144 nmol · L^-1^, Figure 2). [25(OH)D] levels did not differ between participants with different health status (i.e. with or without COPD diagnosis). In the vitamin D_3_ arm, the study was initiated by 14 days of high-dosage vitamin D_3_ intake (10 000 IU^-day^), which led to 42 nmol L^-1^ increases in [25(OH)D] (to 122 ± 24 nmol · L^-1^; range = 82-175 nmol · L^-1^; p < 0.001), with no change in the placebo arm (79 ± 31 nmol · L^-1^; range = 36-167 nmol · L^-1^) (Figure 2). During the remainder of the study (weeks 3-30), vitamin D was ingested at 2 000 IU^-day^, which led to stabilization of [25(OH)D] at elevated levels compared to the placebo arm (Week 13, Δ45 nmol · L^-1^; Week 17, Δ49 nmol · L^-1^; Week 29, Δ46 nmol · L^-1^; Figure 2), resembling the efficacy of previous studies with comparable study protocols (∼2 500 IU^-day^).^87,88^ Conversely, in the placebo arm, [25(OH)D] either declined or was similar to pre-RCT levels (Week 13, −8 nmol · L^-1^; Week 17, −11 nmol · L^-1^; Week 29, −6 nmol · L^-1^; Figure 2), corroborating with changes typically seen in Northern populations during winter months,^27^ with the notable observation that values were slightly higher than expected.^28^ At the onset of introduction to training (Week 13) and throughout the training intervention (Week 17, Week 29), participants in the vitamin D_3_ arm were all vitamin D-sufficient, as classified by the National Academy of Medicine (> 50 nmol · L^-1^),^27^ while in the placebo arm, 13 (Week 13), 12 (Week 17) and 5 (Week 29) participants were vitamin D-insufficient. In both supplementation arms, calcium was ingested at 500 mg · day^-1^ throughout the intervention. Despite this, no changes were seen in calcium or albumin-corrected calcium levels in blood at any time point (Table S8). Levels of the parathyroid hormone decreased throughout the intervention (p = 0.035; Table S8), most likely as an autoregulatory response to increased calcium intake.^89^ Vitamin D_3_ supplementation did not alter this response. Compliance to the supplementation protocol was high in both intervention arms (vitamin D_3_, 99.3 %; placebo, 99.3 %; p = 0.998). Together, these observations suggest that vitamin D_3_ supplementation led to improved vitamin D-status during the intervention, measured as 25(OH)D, whereas placebo led to reduced or maintained levels, with approximately 1/3^rd^ of placebo-receiving participants showing levels associated with impaired muscle functionality at the onset of resistance training.^21,22,90^

### Effects of 12 weeks of vitamin D_3_-supplementation only (weeks 1-12) on muscle strength, performance and characteristics

The main purpose of the initial 12 weeks of vitamin D_3_ supplementation-only was to ensure physiologically elevated [25(OH)D] for a prolonged period of time prior to resistance training, potentially priming muscle cells for plasticity. Vitamin D_3_ supplementation itself had no effect on upper- and lower-body muscle strength and performance, muscle fibre area and characteristics (*m. vastus lateralis*), or hormone concentrations in blood compared to placebo (Figure S1 and Table S2), showing no interaction with health status. Surprisingly, the only exception was 1RM knee extension, for which vitamin D_3_ led to negative changes compared to placebo (Δ-8.4 %; p = 0.008), opposing the seemingly accepted dogma that vitamin D supplementation exerts positive effects on leg muscle strength.^35,91^ Notably, for all muscle strength and muscular performance variables, the initial 12 weeks of supplementation was associated with improved performance when combining the supplementation arms, ranging from 5-71 % (1RM knee extension, 5 %; 1RM chest press 8 %; muscular performance knee extension, 13 %; muscular performance chest press, 71 %; Figure S1, p < 0.05), the only exception being handgrip strength (2 %, p = 0.805). These improvements occurred without any apparent changes in muscle cell characteristics in thigh muscle, including muscle fibre CSA (type I, 4 %, p = 0.573; type II, 9 %, p = 0.312), muscle fibre type proportions (Figure S1; p = 0.127 – 0.901), and total RNA/rRNA expression (p = 0.604 – 1.000). They were hence likely caused by technical, psychological and neural learning effects,^45^ effectuated by repeated exposure to testing prior to and during the supplementation period (Figure S1), as is typically seen in older subjects.^92^ Indeed, dynamic exercises like knee extension and chest press are associated with lower intra-rater reliability than the grip strength test,^45^ which remains unaffected by test-retest,^45^ as was likely the case in the present study.

Overall, the 12-weeks supplementation-only period leading up to the resistance-training period did not lead to any marked changes in mRNA transcriptome profiles. Vitamin D_3_ supplementation was, however, associated with differential changes in the expression of a selection of genes compared to placebo; 25 genes ↑ and 29 genes ↓ (Figure 9a). These included increased expression of B-cell lymphoma 6 and prolyl 4-hydroxylase subunit alpha-1 (BCL6 and P4HA1; Figure 9a), and decreased expression of angiopoietin-like protein 4 (ANGPTL4; Figure 9a). BCL6 and P4HA1 are known to counteract accumulation of reactive oxygen species (ROS),^93–95^ while the level of ANGPTL4 expression is closely correlated with levels of mitochondrial respiration.^96^ These indications were reaffirmed by gene enrichment analyses, which showed less expression of mitochondrial gene sets in the vitamin D_3_ arm (Figure 9b and Table S7). Speculatively, as vitamin D is known to counteract ROS and mitochondrial oxidative stress,^97^ the negative effects of vitamin D_3_ supplementation on the expression of mitochondrial genes may be due to decreased mitochondrial turnover.

### Introductory observations on the quality and general efficacy of the resistance training protocol (weeks 13-28)

Before assessing the effects of combined vitamin D_3_ supplementation and resistance training, it is vital to reaffirm that the protocols and methods held sufficient validity and reliability, including a general assessment of the efficacy of the resistance training intervention. All training sessions were supervised by qualified personnel, as suggested by others,^47^ which likely contributed to the very low drop-out rate (n = 4 during the training period, ∼5 %, Table 1), and ensured high adherence to the protocol (98 %, range 81-100 %, Table 1) and appropriate training progression throughout the intervention (Figure 2). Training volume (repetitions. kg) increased by 20 % (knee extension) and 30 % (leg press) from Week 14 (the first week of training) to Week 18 (the 4th week of training), by 48 % and 54 % to Week 22 (the 8th week of training) and by 65 % and 68 % to Week 27 (the last week of training) (Figure 2). This resembles or exceeds training progression seen in similar studies on previously untrained participants,^98,99^ and was accompanied by progressive increases in perceived exercise intensities (using the Borg RPE-scale^(100)^) (Figure 2). For these training characteristics, no differences were observed between supplementation arms (p = 0.897 - 0.980). The arguably successful completion of the resistance training intervention was accompanied by marked functional and biological adaptations in the participants, including increased muscle strength and performance (e.g. 22 % and 72 % increases in 1RM and muscular performance in knee extension, respectively, p < 0.05, Figure S1), increased muscle mass (e.g. 16 - 24 % increases in muscle fibre CSA for *m. vastus lateralis*, p < 0.05, Figure S1), increases in myonuclei number *per* fibre (30 – 37 %, p < 0.05, Figure S1), alterations in muscle fibre proportions (e.g. type IIX fibre proportions changed from 10 % to 7 %, p < 0.05, Figure S1), and robust alterations in muscle transcriptome profiles (521 and 336 differentially expressed genes at post-introduction resistance training and post-RCT, compared to pre-introduction to resistance training, Figure 9c-d). Importantly, neither of these muscle fibre characteristics changed from pre-RCT to before onset of resistance training (Week 13), suggesting that muscle biopsies sampled before and after the supplement-only period could be regarded as a sampling-resampling event (Figure S1). For muscle strength, the intervention had relative efficiencies of 0.86 % (knee extension) and 1.43 % (leg press) increase per session, which resemble or exceeds expectations based on previous studies of untrained older adults (0.5 – 1.0 % per session).^101–103^

### Analytical measures to increase the validity of vitamin D_3_-based analyses

To ensure valid analyses of the effects of vitamin D_3_ supplementation on muscle-related features, we deemed two precautionary measures to be necessary. First, for muscle strength and muscle performance (apparatus exercises), we defined baseline levels to be equivalent to values collected after 3.5 weeks of introduction to resistance training (main analyses, Figure 2), rather than values collected before its onset, as noted in the preregistration of the study (NCT02598830). At this time point, initial adaptations to training were likely to have occurred, preferably non-hypertrophic effects relating to technical, psychological and neural learning effects,^45^ phenomena that are particularly prominent in older subjects.^92^ Using this time point as baseline arguably strengthens the association between changes in muscle strength and muscle mass, which was the main perspective of our vitamin D_3_-based analyses.

To further minimize the confounding effects of non-hypertrophic increases in strength and performance, all participants conducted a series of repeated tests prior to baseline tests, including five repeated 1RM and muscular performance tests in knee extension and chest press (Figure S1; a, b, e and f), four of which was conducted prior to onset of introduction to training. As expected, this led to progressive increases in strength/performance levels compared to pre-RCT values (defined as the values achieved during the second test, as the first test was conducted at ∼95 % of maximal effort), evident as 4 % (test #4, after 8 weeks of supplementation), 8 % (test #5, pre-introduction to resistance training) and 14 % (test #6, post-introduction to resistance training/baseline) increases in 1RM knee extension (p < 0.05) and 3 % - 5 % - 13 % increases in 1RM chest press, respectively (Figure S1 a and b; notably the third test was conducted at ∼95 % of maximal effort and was omitted from these analyses). These improvements occurred without any apparent hypertrophy in *m. vastus lateralis* of the dominant leg, measured as muscle fibre CSA (pre-RCT vs. pre-introduction to resistance training; type I, p = 0.573; type II, p = 0.312), as previously presented (Figure S1 g), strengthening the notion that the improvements were due to other factors. After adopting the post-introduction-to-training time point as baseline for the strength outcome measures, the efficiency of the intervention on muscle strength was still somewhat higher than expected based on previous observations^101–103^ (1RM knee extension, 0.8 % · session^-1^; 1RM leg press, 1.3 % · session^-1^). Notably, albeit these studies applied less extensive measures to ensure reproducibility, they reported low test-retest variability, which does not concur with our results.^101–103^ For leg press, three tests were performed prior to the baseline test, resulting in similar improvements as observed for knee extension and chest press (14 %). Notably, for other outcome measures, such as muscle biological data, muscle thickness, body composition and endurance-related outcome measures, baseline was defined as onset of introduction to resistance training (Figure 2, Week 13). For variables such as self-reported health, blood variables and lung function, baseline was defined as pre-RCT values (Figure 1; Week −1).

Second, for analyses of the effects of vitamin D_3_ supplementation on changes in muscle mass, we found it necessary to reconsider our choice of using muscle fibre CSA in *m. vastus lateralis* as the primary objective of the study. These data were associated with large degrees of sampling-to-resampling variation, as evaluated using repeated muscle biopsies from the dominant leg, sampled at weeks −1 and 13, i.e. prior to the introduction to resistance training (Figure S2). Rough analyses suggested that we would have needed > 250 fibres of each fibre type to achieve reliable assessment of CSA and > 600 fibres to achieve reliable assessment of fibre type proportions, of which our material contained an average of 118 ± 64/137 ± 69 fibres (type I/type II, range 0 – 428/11 - 424) and 462 ± 265 fibres (range 26 - 1982), respectively (Figure S2). Similar issues have been previously reported for such analyses,^104^ though not in all studies.^105,106^ Speculatively, older adults (with or without COPD) may display larger spatial heterogeneity in muscle fibre characteristics compared to young adults, possibly relating to the age-related remodeling of motor units.^107^ This may have contributed to the high heterogeneity. Despite these inherent issues with muscle fibre analyses, the data provided sufficient resolution to disclose marked increases in muscle fibre CSA and changes in muscle fibre proportions over the entirety of the training intervention, as previously presented (Figure S1).

In order to achieve reliable assessment of changes in muscle mass, we thus had to take on a different approach. Instead of relying on muscle fibre CSA data alone, we developed a combined muscle mass factor, in which change scores from a collection of muscle mass-related outcome measures were combined in a weighted manner (Table S4). This factor included data on muscle fibre CSA, leg lean mass (DXA) and muscle thickness (*m. rectus femoris, m. vastus lateralis*; ultrasound), all of which are known to correlate,^108–110^ and was calculated to accommodate methodological limitation inherent to each of them separately. Corroborating with this, each of these measures separately correlated with the computed muscle mass factor at baseline (r = 0.644 - 0.914) and change scores for each of these measures at post-RCT correlated with the change score for the muscle mass factor (r = 0.331 – 0.939, suggesting the feasibility of our approach; Table S4).The computed muscle mass factor displayed a significant change from baseline to post-RCT (9 %, p < 0.001, Table S4). Following this logic, we also developed similar combined factors for other core outcome domains, including maximal muscle strength and one-legged and whole-body endurance performance (Table S4). For each domain, these factors are reported as the main outcome measure of the study, though separate variables are also presented.

### Effects of vitamin D_3_ supplementation on training-associated changes in maximal muscle strength and lower-limb muscle mass

Participants in both vitamin D_3_ and placebo arms showed increases in every measure of muscle strength and mass, assessed from baseline to after finalization of the resistance training intervention (Figures 4 and 5). These beneficial effects included 12-25 % increases in 1RM muscle strength in legs and upper limbs (Figure 4), 6-11 % increases in muscle torque in legs (Figure 4), 7-26 % increases in muscle fibre CSA and muscle thickness in legs (Figure 5), and 1-3 % increases for lean mass in legs (Figure 5). Unsurprisingly, the combination of these measures into weighted muscle strength and muscle mass factors led to similarly increases over the course of the intervention (13 % ± 8 % and 9 % ± 8 %, respectively; Figures 4 and 5), which was also the case for a calculated score of relative muscle quality (Δmuscle strength factor / Δmuscle mass factor; 4 % ± 10 %, Figure 5).

**Figure 4.**
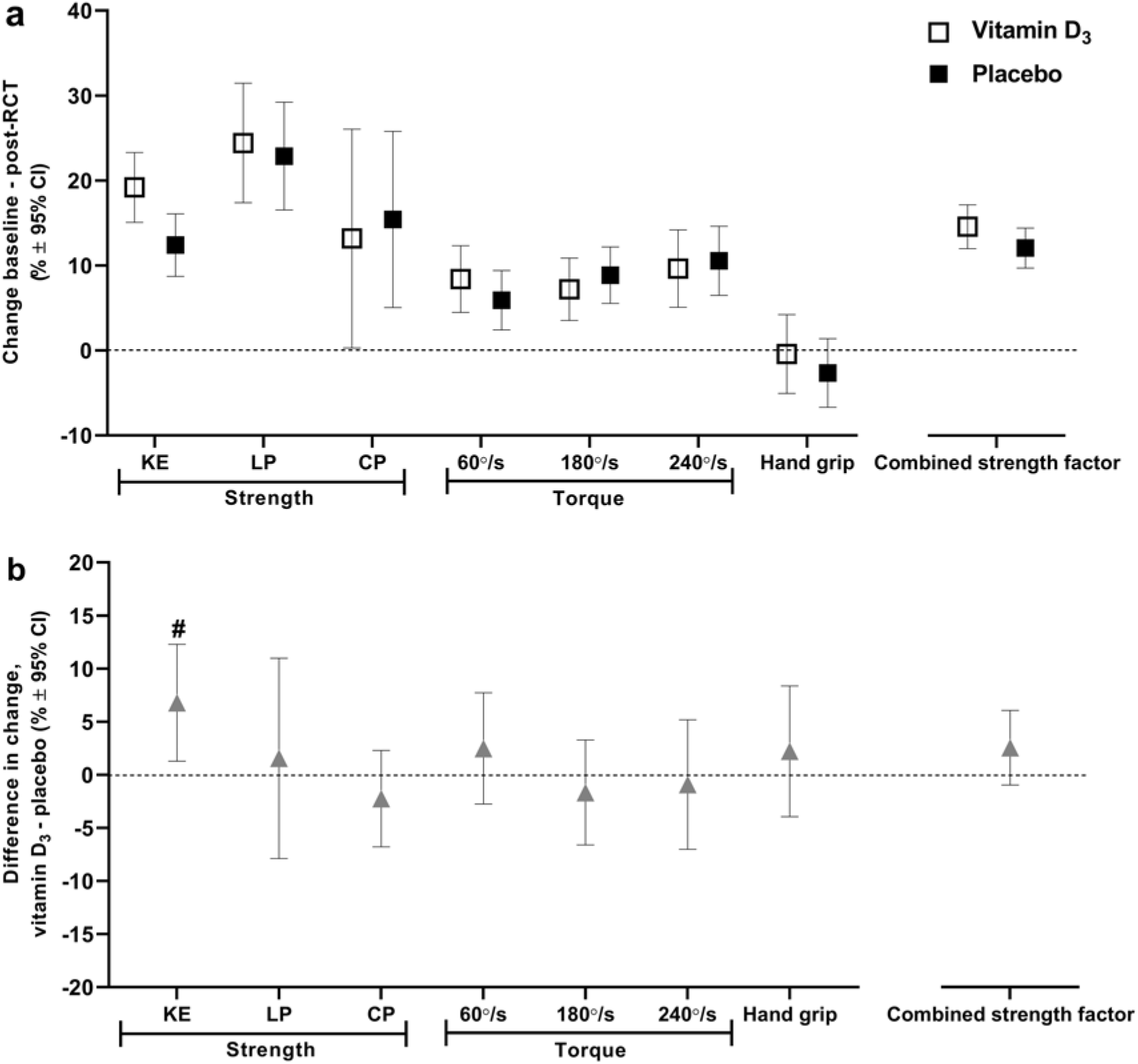
Effects of combined vitamin D_3_ supplementation and resistance training on maximal muscle strength in old adults. Changes in muscle strength from baseline (after three weeks of introduction to resistance training) to post-RCT (a), and differences in changes between vitamin D_3_ and placebo arms (b). KE, one-legged knee extension; LP, one-legged leg press; CP, chest press; maximal torque measured using one-legged knee extension at three velocities; 60, 180 and 240°/second; #, significant difference between vitamin D_3_ and placebo arms; combined strength factor, weighted combined strength factor of unilateral strength measures (one-repetition maximum in KE and LP, and KE torque at 60, 180 and 240°/second). Alpha level at p < 0.05. Data are presented as means with 95 % confidence intervals.

**Figure 5.**
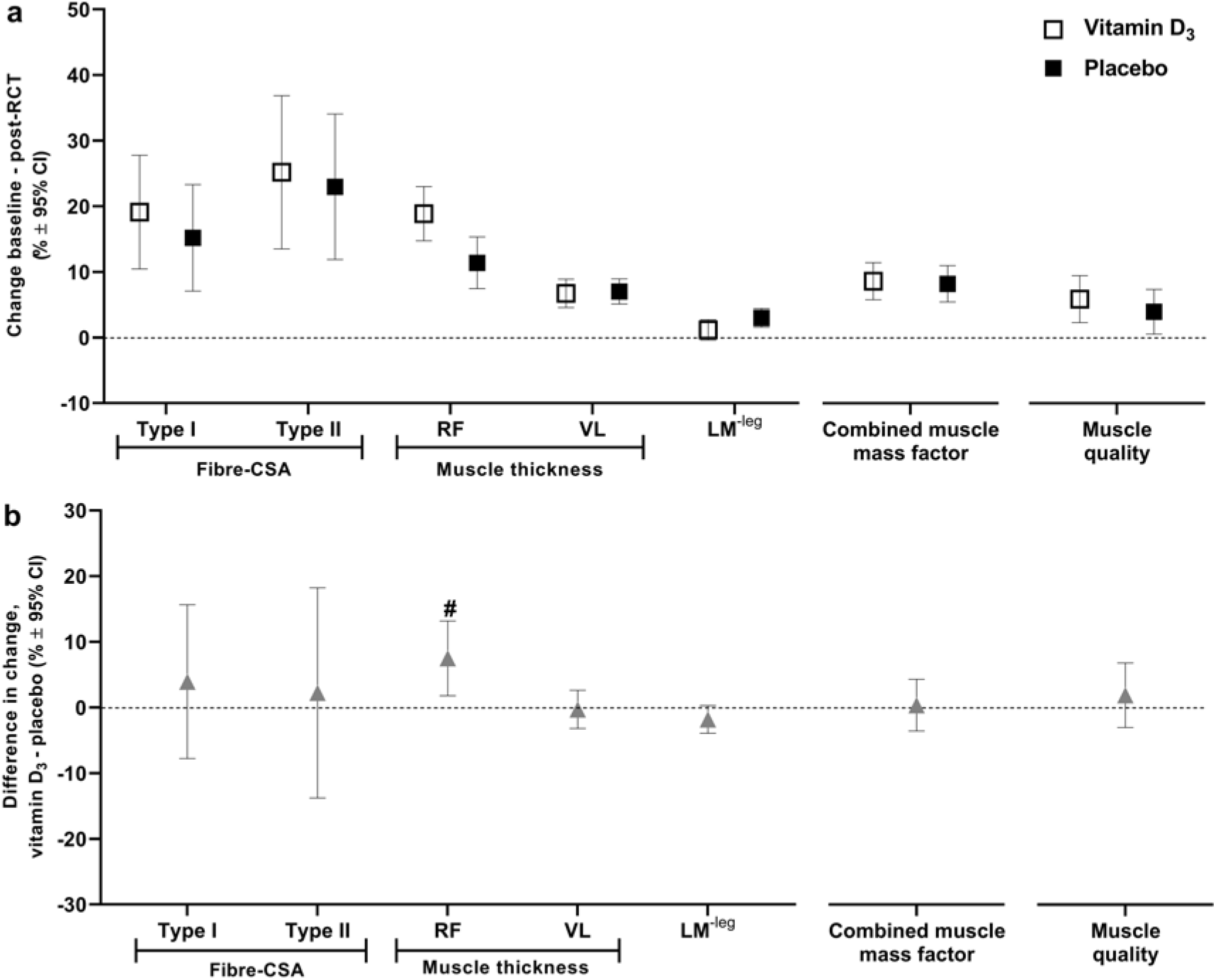
Effects of combined vitamin D_3_ supplementation and resistance training on lower-limb muscle mass in old adults. Changes in lower-limb muscle mass from baseline (before introduction to resistance training) to post-RCT (a), and differences in changes between vitamin D_3_ and placebo arms (b). CSA, cross-sectional area; RF, *m. rectus femoris*; VL, *m. vastus lateralis*; LM^-leg^, leg lean mass *per* leg; #, significant difference between vitamin D_3_ and placebo arms; combined muscle mass factor, weighted combined muscle mass factor including fibre cross-sectional area (type I and type II), muscle thickness (RF and VL) and LM^-leg^; muscle quality, muscle strength factor/muscle mass factor. Alpha level at p < 0.05. Data are presented as means with 95 % confidence intervals.

Overall, vitamin D_3_ supplementation did not affect these outcome measures compared to placebo, primarily assessed as changes in the combined muscle strength and muscle mass factor (strength, Δ2.5 % (95 % CI, −1.0, 6.0), p = 0.194; mass, Δ0.4 % (95 % CI, −3.5, 4.3), p = 0.940, Figures 4 and 5), and secondarily as changes in each of the underlying outcome measures (i.e. seven measures of muscle strength and three measures of muscle mass; Figures 4 and 5). This lack of beneficial effects were also evident for changes in relative muscle quality (Δ1.9 % (95 % CI, −3.0, 6.8), p = 0.415; Figure 5). Vitamin D_3_ supplementation thus had no main effect on training-associated changes in muscle functionality and gross muscle biology in the participants. While this conclusion coheres with the few comparable studies assessing the effect of combined vitamin D_3_ intake and resistance training,^40,42–44^ it contrasts the conclusion drawn in the only available meta-analysis on this subject, wherein vitamin D_3_ supplementation was associated with augmented increases in muscle strength in older adults.^41^ Notably, among the selection of ten specific outcome measures, two did not conform with the main finding. Vitamin D_3_ was associated with beneficial effects for changes in 1RM knee extension (Δ6.8 % (95 % CI, 1.3, 12.3), p = 0.016; Figure 4) and muscle thickness of *m. rectus femoris* (Δ7.5 % (95 % CI, 1.8, 13.2), p = 0.011; Figure 5). For 1RM knee extension, the effect was interrelated with the negative development seen from pre-RCT to pre-introduction to training in the vitamin D_3_ arm (Figure S1). Indeed, when assessing the effect of vitamin D_3_ on 1RM knee extension from pre-to post-RCT (rather than from baseline at post-introduction to training), no beneficial effect was observed compared to placebo (Δ-2 % (95 % CI, −12, 7), p = 0.628; Table S2). As for muscle thickness in *m. rectus femoris*, we did not collect data pre-RCT and can thus not assess if this variable followed the same pattern as 1RM knee extension throughout the intervention. The observed benefits of vitamin D_3_ supplementation for changes in *m. rectus femoris* thickness contrasts observations made for *m. vastus lateralis* thickness (Δ-0.3 %, p = 0.838), and even oppose those made for lean mass of the legs, which tended to increase less in the vitamin D_3_ arm compared to the placebo arm (Δ-1.8 %, p = 0.090).

So far, our analyses have focused on the main effect of vitamin D_3_ supplementation on training-induced development of muscle strength and mass, and have thus neglected potential interactions with other independent variables such as pre-RCT levels of 25(OH)D, health status (COPD vs non-COPD) or training modality (high-load, 10RM, vs low-load, 30RM). The benefits of vitamin D_3_ supplementation were expected to be more pronounced in participants with low baseline levels of 25(OH)D. This hypothesis was based on observations made in cohort studies, wherein subjects with levels < 30-50 mmol. L^-1^ are more likely to show adverse muscle phenotypes.^21–23^ To investigate this perspective, participants in each supplementation arm were divided into quartiles based on pre-RCT 25(OH)D levels in blood (Figure S3). This resulted in two lower quartiles, one for the vitamin D_3_ arm (vitamin D3_low_, [25(OH)D]_mean_ = 49.5 mmol. L^-1^, n = 8), and one for the placebo arm (placebo_low_, with [25(OH)D]_mean_ = 47.4 mmol. L^-1^, n = 12) (Figure S3). At the onset of introduction to resistance training, 25(OH)D levels in vitamin D3_low_ had increased to 103.3 mmol. L^-1^ (range 76-138), with all participants being classified as sufficient (> 50 nmol. L^-1^),^17^ whereas levels in placebo_low_ remained unchanged (45.5 nmol. L^-1^, range 22-71), with 9 out of 12 participants having insufficient levels (< 50 nmol. L^-1^). Within each of the pre-RCT 25(OH)D quartiles, the effect of vitamin D_3_ and placebo supplementation on training-induced changes in muscle strength and mass (using the combined factors) were assessed. With exception of one quartile (muscle strength factor, quartile 3, p = 0.048; Figure S3), there was no beneficial effect of vitamin D_3_ supplementation in any quartile (e.g. vitamin D3_low_ vs placebo_low_, muscle strength, Δ-2.0 % (95 % CI, −8.0, 3.9, p = 0.496) (Figure S3). Instead, in vitamin D3_low_, changes in muscle mass was lower than in placebo_low_ (Δ-6.5 % (95 % CI, −12.7, −0.27), p = 0.041; Figure S3), suggesting that vitamin D_3_ supplementation may have compromised changes in muscle mass in subjects with low pre-RCT 25(OH)D levels. Adding to this, participants in the different quartiles responded quite similarly to resistance training, irrespective of supplementation arms, evident as no interaction between 25(OH)D quartiles/supplementation arm and changes in muscle strength (p = 0.237) or muscle mass (p = 0.159). Arguably, the statistical power of these analyses were not sufficiently high to conclude on this perspective.

The impact of vitamin D_3_ supplementation on muscle strength and muscle mass factors did not interact with health status (COPD vs non-COPD) or training modality (10RM vs 30RM) (Table S2). However, it should be noted that for selected specific outcome measures, interactions were found with both of these independent variables (summarized in Table S2), including an interaction between changes in type II-fibre CSA and COPD/non-COPD, and between changes in 1RM knee extension/vastus lateralis thickness and 10RM/30RM. In addition to these interaction analyses, we also investigated the potential relation between effects of vitamin D_3_ supplementation and baseline body fat proportions, as overweight and obese have been shown to have decreased bioavailability of vitamin D due to deposition of 25(OH)D in body fat compartments (while concomitantly showing attenuated anabolic response to resistance exercise^(111)^).^112^ As in our quartile-based analyses of the effects of pre-RCT [25(OH)D], we divided each supplementation arm into quartiles based on baseline body fat percentage and body mass index (BMI), resulting in two higher quartiles for each measure of body mass composition, one for each supplementation arm (e.g. quartile_high fat percentage_ vitamin D_3_ arm, mean 40 % ± 4 %, range 37 – 47 %, n = 8; quartile_high fat percentage_ placebo arm, mean 46 % ± 6 %, range 40 – 57 %, n = 9). Analyses did not reveal an effect of baseline body fat proportions for changes in [25(OH)D] (fat percentage, p = 0.432; BMI, p = 0.369) or muscle mass factor (fat percentage, p = 0.355; BMI, p = 0.293) (Figure S4). However, it did have an effect on changes in the muscle strength factor (fat percentage, p = 0.016; BMI, p = 0.706), i.e. in quartile_high fat percentage_, vitamin D_3_ supplementation was associated with larger increases in muscle strength compared to placebo (fat percentage, Δ 5.8 % (95 % CI, 0.5, 11.0), p = 0.032; BMI, Δ7.8 % (95 % CI, 2.5, 13.1), p = 0.005; Figure S4 and Table S2), suggesting beneficial effects of vitamin D_3_ supplementations in subjects with high proportions of body fat, opposing our initial expectations.

### Effects of vitamin D_3_ supplementation on training-associated changes in one-legged and whole-body endurance performance

Participants in both vitamin D_3_ and placebo arms showed improvements in one-legged and whole-body endurance performance over the course of the resistance training intervention (Figure 6). These beneficial effects included 42 - 74 % increases in one-legged muscular performance (repetitions to exhaustion at 50 % of 1RM pre-RCT in knee extension and chest press, Figure 6), 7 - 9 % increases in peak power output (W_max_) in one- and two-legged cycling (Figure 6), 3 - 5 % reductions in O_2_ costs of submaximal one-legged cycling (Table S2), and 6 - 10 % increases in functional performance (sit-to-stand test and 6-min step test, Figure 6). In accordance with this, we observed marked increases in weighted one-legged and whole-body endurance performance factors (one-legged, vitamin D_3_ 25 % ± 19 %, placebo 22 % ± 11 %; whole-body, vitamin D_3_ 9 % ± 8 %, placebo 7 % ± 6 %; Figure 6), calculated from change scores of relevant specific outcome measures (excluding O_2_-costs of one-legged cycling). These effects cohere well with previously observed effects of resistance training on endurance variables in older adults.^113–115^

**Figure 6.**
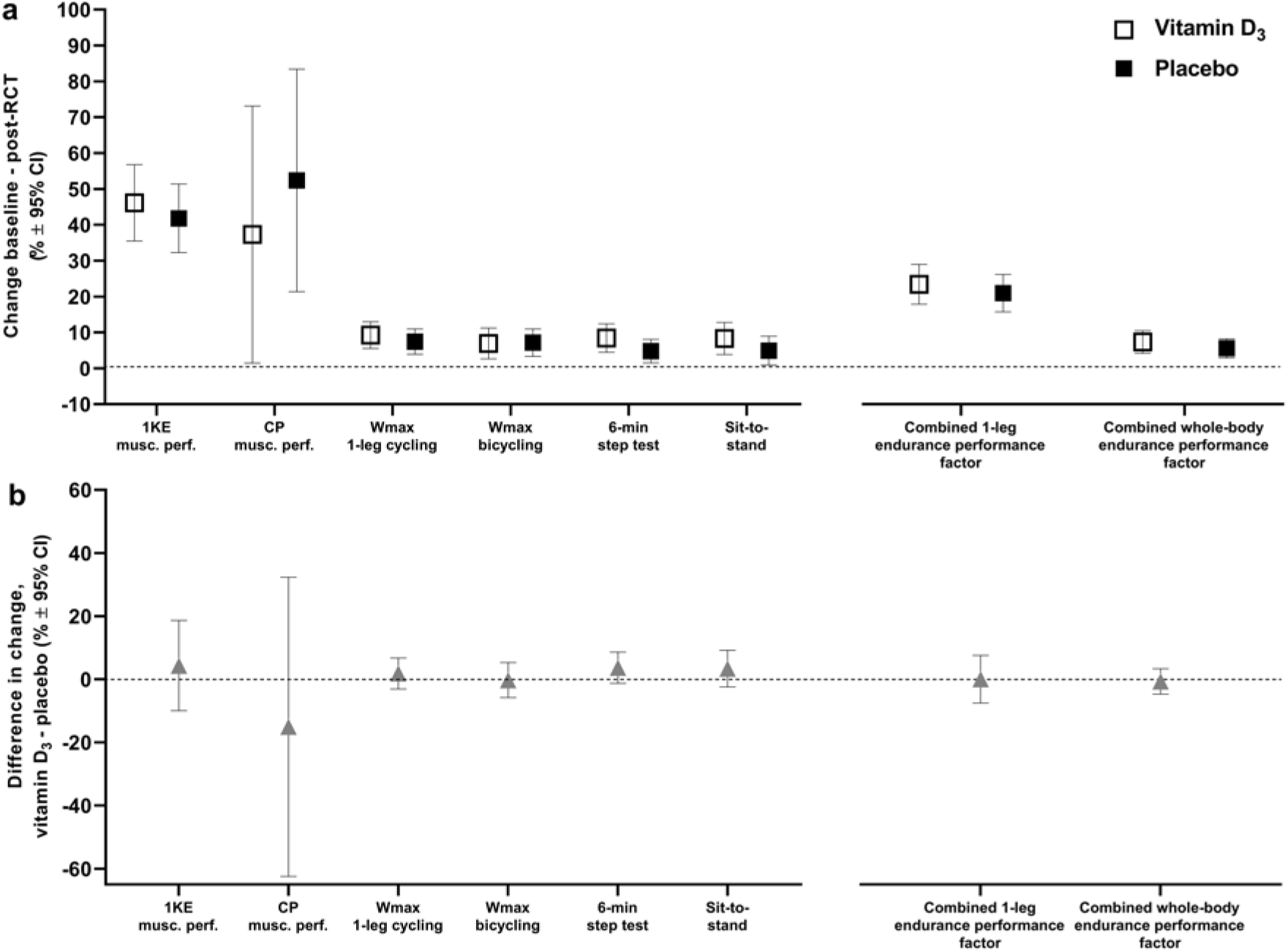
Effects of combined vitamin D_3_ supplementation and resistance training on one-legged and whole-body endurance performance in old adults. Changes in endurance performance from baseline (before introduction to resistance training) to post-RCT (a), and differences in changes between vitamin D_3_ and placebo arms (b). 1KE, repetitions to failure in one-legged knee extension (50 % of pre-intervention 1RM); CP, repetitions to failure in chest press (50 % of pre-intervention 1RM); Wmax, maximal power output; 6-min step test, maximal number of steps achieved during six minutes; Sit-to-stand, maximal number of sit-to-stands achieved during one minute; combined 1-leg endurance performance factor, weighted combined one-legged endurance factor including 1KE muscular performance and one-legged cycling Wmax; weighted combined whole-body endurance factor including Wmax bicycling, 6-min step test and sit-to-stand test. Alpha level at p < 0.05. Data are presented as means with 95 % confidence intervals.

Vitamin D_3_ supplementation had no effect for any of these outcome measures compared to placebo, neither for weighted endurance performance factors (one-legged, Δ2 % (95 % CI, −5, 10), p = 0.773; two-legged, Δ2 % (95 % CI, −2, 6), p = 0.636; Figure 6), nor for any of the specific outcome measures (Figure 6). For one-legged and whole-body endurance factors, there was no interaction between baseline 25(OH)D quartiles and effects of vitamin D_3_ supplementation (one-legged, p = 0.950; whole-body, p = 0.266; Figure S3 and Table S2), nor was there any interactions with health status (one-legged, p = 0.747, whole-body, p = 0.129, Table S2) or training modality (one-legged, p = 0.719, Table S2).

### Effects of vitamin D_3_ supplementation on training-associated changes in muscle fibre characteristics and transcriptomics

Participants in both vitamin D_3_ and placebo arms showed marked changes in muscle fibre characteristics over the course of the training intervention. These included decreased type IIX muscle fibre proportions from 10 % to 7 % (Figure 7), increased type IIA proportions from 26 % to 29 % (Figure 7), increased type IIA/IIX hybrid fibres abundances from 2.6 % to 3.2 % (Table S2), and 25 - 48 % increases in myonuclei number *per* muscle fibre (Figure 7). Changes in IIX and IIA proportions were verified using qPCR, showing decreased levels of type IIX mRNA abundance and increased levels of type IIA (Figure 7), calculated using the gene family-profiling approach.^76^. In addition, qPCR analyses revealed increased type I mRNA proportions after the training intervention (Figure 7). These observed changes in muscle fibre-type characteristics corroborate well with previous studies in older adults,^116–118^ though increases in myonuclei number per muscle fibre are not consistently reported.^119^ Vitamin D_3_ supplementation had no effect on training-associated changes in muscle fibre proportions or myonuclei content compared to placebo (Figure 7).

**Figure 7.**
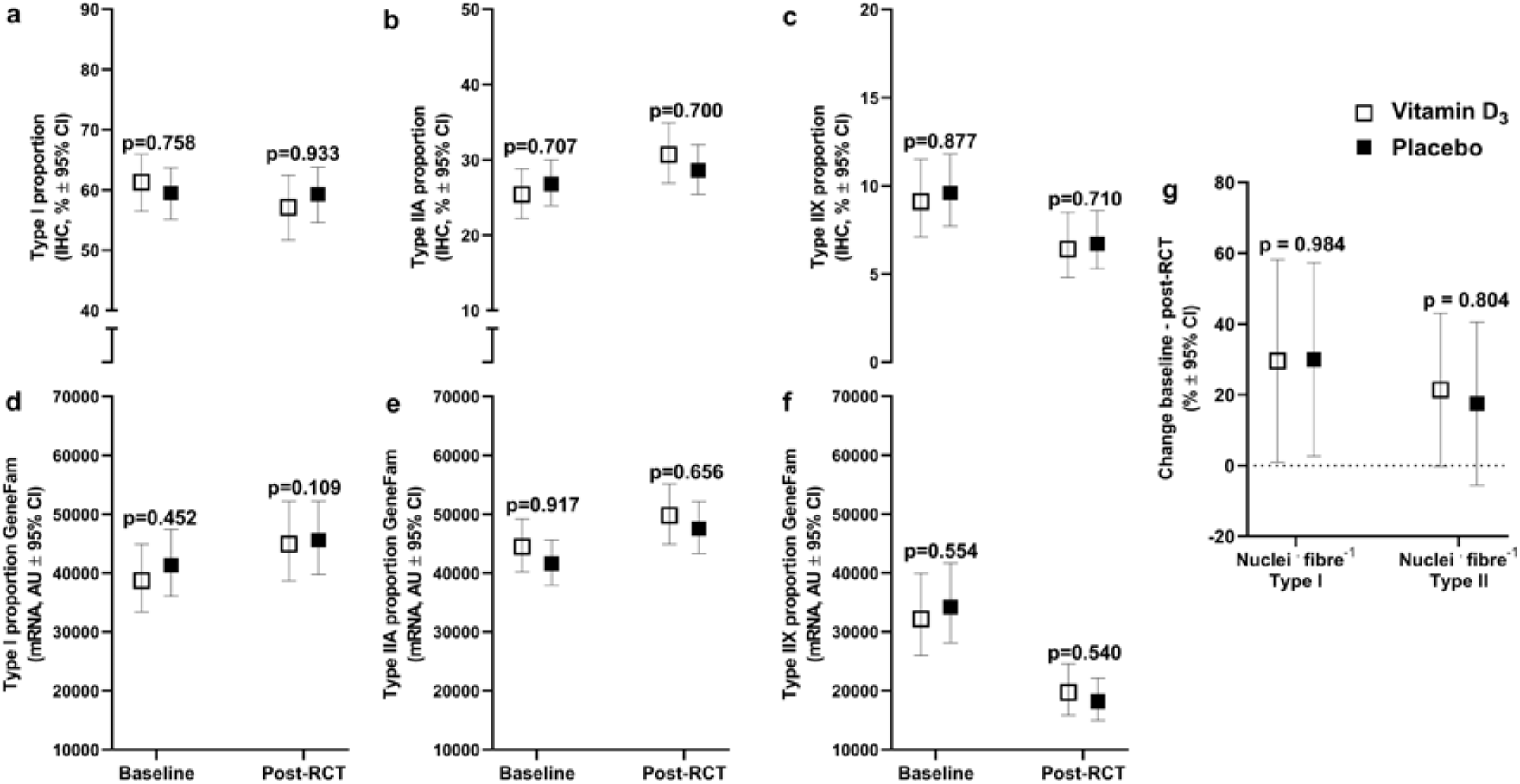
Effects of combined vitamin D_3_ supplementation and resistance training on muscle fibre type proportions and myonuclei per fibre in *m. vastus lateralis* of old adults. Muscle fibre type proportions (a-f) at baseline (before introduction to resistance training) and post-RCT measured using immunohistochemistry (a-c) and qPCR (gene family profiling (GeneFam)-normalized myosin heavy chain mRNA expression, d-f), and changes in myonuclei count per type I and type II fibre from baseline to post-RCT (g). Significant changes were observed for fibre type IIA and IIX using both methods (significant increase and decrease, respectively; p < 0.05). For fibre type I, an increased expression was present using qPCR (p < 0.05), but no change was observed for immunohistochemistry (p = 0.322). P-values denotes the statistical difference between the supplementation arms. RT, resistance training. Data are presented as means with 95 % confidence intervals.

The training intervention resulted in 1.14 – 1.16 fold increases in total RNA per unit tissue muscle weight (Figure 8), a proxy marker for ribosomal RNA content that has previously been associated with training-induced changes in muscle growth and strength.^67,120^ Similar increases were found for the mature ribosomal species 18s (1.18 fold) and 28s (1.16 fold), in addition to the 45s pre-ribosomal rRNA (1.19 fold) using qPCR (Figure 8). No changes were observed for 5.8s (1.07 fold, p = 0.722) or 5s (1.06, p = 0.940) following the entire training intervention. Notably, for analyses of total RNA and ribosomal RNA, we included an additional time point in our main analyses, i.e. in muscle biopsies sampled after introduction to training (3.5 weeks, 7 sessions), as early increases in total RNA seem to associate with long-term chronic responses to training, making it a potential hallmark of muscle plasticity.^67^ As expected, 3.5 weeks of training led to marked increases in total RNA (1.10 – 1.21 fold) and all ribosomal RNA species studied (1.13 – 1.27 fold) (Figure 8). The changes in total RNA and rRNA expression corroborates quite well with changes observed in healthy, young subjects,^67^ although with a slightly reduced impact. However, this does not comply with the previous finding of no change in RNA:DNA following resistance training in older subjects.^121^ Vitamin D supplementation had no effect on training-associated changes in total RNA or rRNA expression compared to placebo.

**Figure 8.**
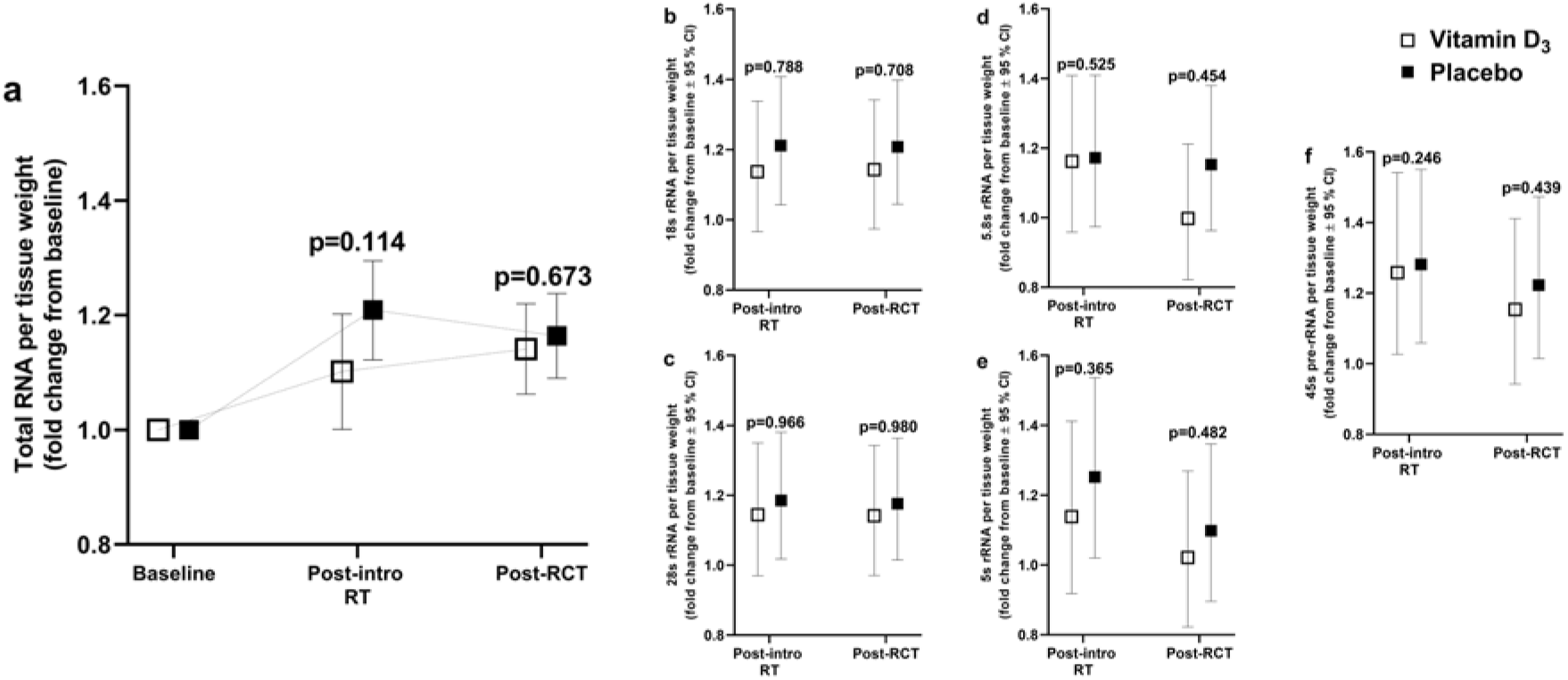
Effects of combined vitamin D_3_ supplementation and resistance training on total RNA abundances and rRNA expression in *m. vastus lateralis* of old adults. Total RNA (a), 18s rRNA (b), 28s rRNA (c), 5.8s rRNA (d), 5s rRNA (e), and 45s pre-rRNA (f) abundances at baseline (before introduction to resistance training) and post-RCT. Significant increases from baseline – post-introduction to resistance training were present for all variables (p < 0.05). From baseline – post-RCT significant increases were present for all variables (p < 0.05), with the exception of 5.8s rRNA (p = 0.722) and 5s rRNA (p = 0.940). RT, resistance training. P-values denotes the statistical difference between the supplementation arms. Alpha level at p < 0.05. Data are presented relative to amounts of tissue weight. Data are presented as means with 95 % confidence intervals

The training intervention led to marked changes in muscle mRNA transcriptome profiles, with 521 genes being differentially expressed (DE) after 3.5 weeks of resistance training (456 genes ↑, 65 genes ↓, compared to pre-introduction to resistance training) and 336 genes being DE after 13 weeks of resistance training (post-RCT; 275 genes ↑, 61 genes ↓) (Figure 9c and 9d; the two supplementations arms were combined in these analyses). In effective library size-based analyses, GO enrichment analyses revealed increased expression of gene sets relating to extracellular matrix after 3.5 and 13 weeks of resistance training (Figure 9e), which coheres well with previous studies.^122^ Moreover, increased expression of gene sets involved in inflammatory response and leukocyte migration were present after 3.5 weeks of resistance training, and increased expression of gene sets involved in blood vessel morphogenesis were present after 13 weeks of resistance training (Figure 9e presents gene sets for extracellular matrix and blood vessel morphogenesis; Table S5 presents the most enriched gene sets with resistance training). The enrichment analysis also revealed decreased expression of gene sets involved in ribosomal functions after 3.5 and 13 weeks of resistance training (Figure 9e), potentially contradicting the notion that increased ribosomal biogenesis is a prerequisite for training-associated muscular adaptations.^67,120^ Importantly, this potential negative effect of resistance training on expression of ribosomal genes disappeared after accounting for amounts of muscle tissue used during RNA-seq library preparations (Table S5), indicating that ribosomal genes is not negatively influenced when expressed per unit tissue weight. This was the only major difference between library size and tissue-offset normalization.

Vitamin D_3_ supplementation had no effect on training-associated changes in mRNA transcripts after neither 3.5 weeks (Figure 9f) nor 13 weeks of resistance training (Figure 9g). This suggests that no single gene was affected by combined vitamin D_3_ supplementation and resistance training compared to resistance training-only. However, enrichment analyses showed traces of differential changes in expression between supplementation arms after both 3.5 and 13 weeks of resistance training (Figure 9h). After 3.5 weeks of training, the vitamin D_3_ arm showed increased expression of gene sets involved in muscle cell differentiation, actin cytoskeleton, cell junctions and blood vessel morphogenesis. These initial responses to resistance training should be interpreted with caution, as they were only evident in GSEA analyses and not in rank-based analyses (Figure 9h and Table S6). After 13 weeks of resistance training, the vitamin D_3_ arm showed increased expression of gene sets involved in endothelial function and blood vessel morphogenesis compared to placebo (Figure 9h; regulation of leading-edge genes in the GO gene set “blood vessel morphogenesis” are illustratively denoted in Figure 9g and the 10 most affected genes are presented in Figure 9i). This agrees with previous observations of a positive relationship between 25(OH)D-status and endothelial function, possibly interacting with the predominant endothelium-derived vasodilator, nitric oxide.^97^ Indeed, it coheres well with a recent study, which showed favorable effects of combined vitamin D_3_ supplementation and resistance training on flow-mediated dilation of blood vessels and blood pressure in postmenopausal women.^123^ Unfortunately, endothelial function was not assessed in the current study.

**Figure 9.**
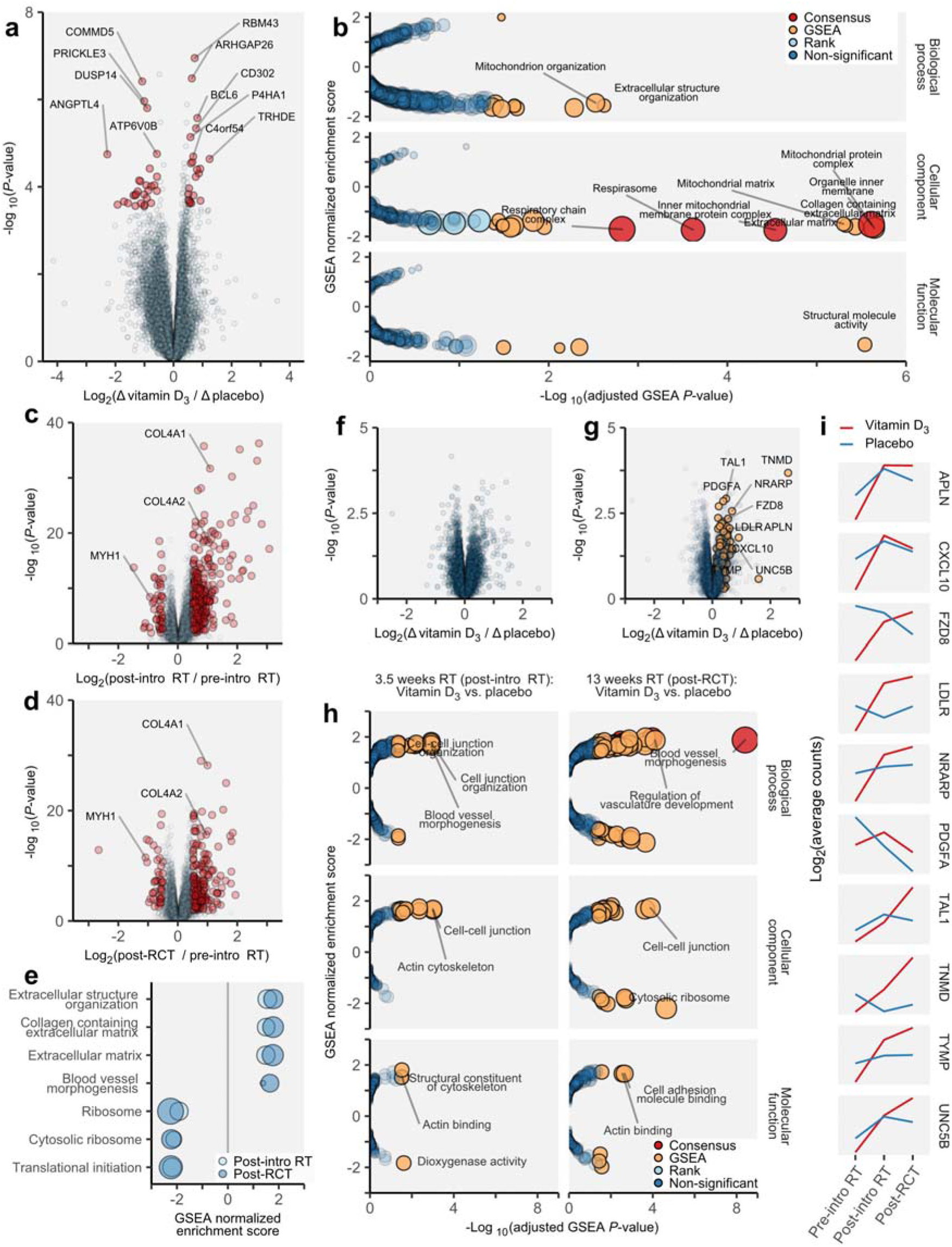
Effects of 12 weeks of vitamin D_3_ supplementation-only (a-b), 3.5/13 weeks of resistance training-only (c-e) and 3.5/13 weeks of combined vitamin D_3_ supplementation and resistance training (f-i) on mRNA transcriptome profiles in *m. vastus lateralis* of old adults. After 12 weeks of supplementation-only, numerous genes were differentially expressed (DE) between the vitamin D_3_ and the placebo arm (a; Δ, pre-introduction to resistance training / pre-RCT). Gene ontology (GO) enrichment analyses showed that these genes were primarily related to mitochondrial function (b; positive/negative GSEA-normalized enrichment scores indicates higher/lower expression of gene sets in the vitamin D_3_ arm compared to the placebo arm). Resistance training-only led to robust changes in gene expression at both 3.5 weeks (c; post-intro resistance training – pre-intro resistance training) and 13 weeks (d; post-RCT – pre-intro resistance training), including increased expression of collagen type IV α1 and α2 genes (COL4A1 and COL4A2, respectively) and decreased expression of the myosin heavy chain IIX gene (MYH1). The three most enriched gene sets with increased and decreased expression, in addition to the “blood vessel morphogenesis” gene set are shown in e (light blue, 3.5 weeks; dark blue, 13 weeks; according to the GSEA enrichment score). Combined vitamin D_3_ supplementation and resistance training did not lead to differential changes in expression for a singular gene compared to placebo at neither 3.5 weeks (f; Δ, post-introduction to resistance training / pre-introduction to resistance training) nor 13 weeks of resistance training (g; Δ, post-RCT / pre-introduction to resistance training; orange dots/genes denotes leading edge genes from the “blood vessel morphogenesis” GO gene set, i.e. the most highly enriched gene set between supplementation arms after 13 weeks of resistance training). GO enrichment analyses of differentially regulated gene sets between the vitamin D_3_ and the placebo arms following 3.5 weeks (left panel, h) and 13 weeks of resistance training (right panel, h; positive/negative GSEA-normalized enrichment scores indicates higher/lower expression of gene sets in the vitamin D_3_ arm compared to the placebo arm). i) Timeline for the 10 most affected genes between vitamin D_3_ and placebo arms belonging to the “blood vessel morphogenesis” GO gene set. RT, resistance training; Consensus, when both the non-directional rank-based enrichment test and the directional gene-set enrichment analysis (GSEA) turned out significant. In Figure 8b, e and h, circle sizes of gene sets are relative to p-values, i.e. larger circles indicate lower p-values (See Table S5-7 for exact p-values).

### Effects of vitamin D_3_ on hormones in blood and health-related outcome measures

In general, the intervention was associated with beneficial changes for several health-related variables, including reduced levels of lipids (triglycerides and low-density lipoprotein/LDL), reduced levels of fat mass (total and visceral fat) and improved self-reported health (Table S8). Conversely, a small but undesirable decrease was observed in lung capacity, measured as forced ventilatory capacity (FVC) (Table S2). The intervention was not associated with changes in whole-body bone mineral density or changes in serum levels of hormones, except for decreased levels of parathyroid hormone (Table S8), as previously presented. For most of the health variables, there was no effect of vitamin D_3_ supplementation (Table S2 and S8), with exception of cortisol levels in blood, which increased more in the vitamin D_3_ arm (Table S8), and lung function measured as FEV_1_/FVC-ratios, which declined in subjects with COPD in the vitamin D_3_ arm (Table S2).

General health. For the general health benefits of the intervention, the −6 % reductions in triglyceride levels and the −4 % reductions of LDL in serum are of particular interest (with no changes being observed for HDL) (Table S8). This lowered the number of participants with diagnostically elevated LDL levels (≥ 4.1 mmol. L^-1^)^(124)^ from 17 to 13, emphasizing the potential benefits of resistance training for lipid profiles, as has previously been shown to be equivocal.^125,126^ This was accompanied by −2.7 % reductions in whole-body fat mass and −5.9 % reductions in visceral fat mass (Table S8). The observed reduction in visceral fat mass is noteworthy, as its relative change was 2.1-fold greater than the change in overall fat mass, though they were largely correlated (Pearson’s r = 0.70; p < 0.001). This is not an uncommon observation^(127–129)^ and suggests that resistance training leads to targeted metabolism of visceral fat, potentially triggered by interleukin-6,^127^ which is indeed secreted from skeletal muscle in response to such training.^130^ Overall, these data support the notion that resistance training is an effective strategy for improving long-term cardiovascular health.^125,131^ The intervention was also effective in counteracting the age-related decline in muscle mass, leading to 1.4 % increases in total lean body mass (p < 0.001) (Table S8) and reduced the number of participants that could be defined as sarcopenic from 16 % (11 subjects) to 12 % (8 subjects). Sarcopenia was defined as appendicular skeletal muscle mass (kg)/m^2^ greater than two standard deviations below the sex-specific means of young adults.^61^

#### Steroid hormones

The lack of effects of vitamin D_3_ supplementation on levels of anabolic steroid hormones such as testosterone did not comply with our initial hypothesis, as we presumed a positive association between vitamin D levels and testosterone levels, based on previous observations from vitamin D_3_ supplementation studies^(53)^ and cohort studies.^132^ However, our finding is in line with most vitamin D supplementation studies, reporting no effect on testosterone in blood.^133,134^ In contrast to this, an effect was found of vitamin D_3_ supplementation on serum cortisol levels compared to placebo (Δ48 nmol. L^-1^, p = 0.038; Table S8), though no main effect of time was observed for cortisol levels. While this does not comply with the notion that [cortisol] and [25(OH]D] is inversely related to each other,^135^ the observed relative increase in the vitamin D_3_ arm may instead be a feed-back response to increased calcium fluxes (fortified by the calcium intake), as glucocorticoids such as cortisol are known to oppose the hypercalcemic effects of high levels of vitamin D.^136^ This also offers an explanation to the lack of increases in calcium levels and albumin-corrected calcium levels in serum (Table S8). There is a notable paucity in data on the effects of vitamin D supplementation on changes in cortisol, making our observation difficult to interpret. The vitamin D_3_-effect on cortisol did not interact with health status (p = 0.122) and was not associated with pre-RCT [25(OH)D] quartiles (p = 0.179) or with combined [25(OH)D] quartiles and supplementation arms (p = 0.345).

#### Lung function

The small −1.95 % reduction in FVC seen after the 28 week long RCT (p = 0.006; Table S2) was surprising, as exercise is generally accepted to be beneficial for lung functionality, so also resistance training.^137,138^ As such, the observed decline may not have been caused by negative effects of the intervention per se (including resistance training), but may instead be related to age- and COPD-associated declines, as their numerical value corroborates with those seen in older adults and elderly over a comparable period of time (current study, −2.85 % (COPD, 0.41 %; non-COPD, −3.97 %) vs −2.46 % per year^(139)^). Notably, other measures of lung function, such as forced ventilatory volume in one second (FEV_1_ and predicted FEV_1_) and FEV_1_/FVC, were not affected by the intervention (Table S2). These are also prone to age-related declines.^139^ Still, our data may advocate incorporation of endurance training into lifestyle therapy programs to oppose the negative effects of age and disease on lung function.^137^

The negative effects of vitamin D_3_ on lung function, measured as FEV_1_/FVC (Δ-2.9 %-points, p = 0.012; Table S2), was also surprising. This effect showed a clear interaction with health status, and as such was only evident in COPD patients in the vitamin D_3_ arm, which showed Δ-8.4 % reductions compared to placebo (Table S2). This subgroup-analysis is however clearly weakened by the small sample size (COPD, n = 9 vs n= 11, vitamin D_3_ vs placebo). The negative effect of vitamin D_3_ on FEV_1_/FVC did not interact with pre-RCT levels of FEV_1_/FVC, but surprisingly, in another subgroup-analysis, pre-RCT 25(OH)D vitamin D3_low_ quartile was associated with larger decrement in FEV_1_/FVC than placebo_low_ (Δ-5.4 %-points, p = 0.009). This observation is difficult to explain. It largely opposes the notion that vitamin D deficiency leads to impaired lung functions.^140^ Speculatively, it also contradicts the previously observed benefits of vitamin D supplementation on the occurrence of exacerbations in COPD patient,^141^ though this effect may primarily relate to improved functions of the immune system, rather than improved lung function *per se*. In the current study, only one exacerbation incidence was registered among the COPD participants during the intervention (vitamin D_3_ arm, prior to training period). Unfortunately, we do not have data from a negative control group, and as such cannot compare our findings to a non-intervention outgroup. More research is clearly needed to elucidate on the consequences of resistance training and vitamin D_3_ supplementation for lung functionality.

#### Adverse effects of the intervention

Overall, neither vitamin D_3_ supplementation nor resistance training was associated with adverse effects or events during the intervention, with potential exception of certain aspects of lung function, as previously discussed, and iron biology.

Primarily, a health survey was administered to the participants on a weekly basis. This included rating of 11 potential discomforts relating to digestion problems, sleep problems, issues with the urinary system, issues with the vestibular system and dermal irritations (Table S2). No effect of vitamin D_3_ supplementation was found for any of these variables. In the health survey, participants were also asked to rate their experienced health on a point-scale from 0-10. This self-reported conception of health improved from 6.3 ± 1.6 to 7.1 ± 1.6 (p < 0.001, Table S2), with no difference between supplementation arms (p = 0.433).

The intervention was associated with alterations in serum levels of markers of iron biology. Specifically, serum levels of Fe^2+^ and ferritin decreased (−12 % and −16 %, respectively), while levels of transferrin remained unchanged (−0.4 %) (Table S8). Speculatively, this may have affected biological processes such as hemoglobin production and the oxygen-carrying capacity of blood, which was not measured. However, no traces of such adverse effects were found in maximal oxygen uptake (Table S2), which did not change over the course of the training intervention and is known to be closely correlated with total hemoglobin mass.^142^ The observed alteration in iron biology may have been due to the daily intake of 500 mg calcium in both supplementation arms, which is known to exert negative effects on iron absorption in humans.^143^ The rationale behind including calcium supplementation as part of the study protocol was to ensure sufficient levels of calcium in both supplementation arms, facilitating potential accretion of bone in response to resistance training, particularly so in the vitamin D_3_ arm.

The intervention was not associated with training-associated injuries, with only five (6 %) reporting discomforts with training towards the end of the intervention and only four (5 %) participants leaving the study during the resistance training intervention, neither of which dropped out because of injuries associated with the training. As such, serum levels of markers of muscle tissue damage (creatine kinase and aspartate aminotransferase) actually decreased during the intervention, with no effects of vitamin D_3_ supplementation (Table S8). Notably, these latter analyses may have been affected by pre-RCT testing, as these were performed during the week preceding blood sampling, and may have contributed to increased levels of creatine kinase and aspartate aminotransferase.^144^ Such responses are typically upon frequent conduction of exercise.^145^ Supervised resistance training can safely be advocated for both healthy older adult and persons with COPD.

### Concluding remarks

The study was conducted as a double-blinded RCT, addressing the effects of 12 weeks of vitamin D_3_ supplementation only (in average, 3 333 IU. day^-1^), followed by 13 weeks of combined vitamin D_3_ (2 000 IU. day^-1^) and resistance training on training-associated adaptations in a mixed population of old adults. Vitamin D_3_ supplementation is often hailed as an ergogenic aid for optimizing the outcome of resistance training, and is recommended for a variety of human populations, ranging from healthy subjects to athletes and chronically diseased subjects.^7,20^ Vitamin D is thus presumed to play an important role in training-associated muscle plasticity. Despite this, its importance for humans remains largely elusive, with current knowledge largely stemming from animal research,^56^ and the few existing human studies providing limited, uncertain and contradicting results.^41–44^ Indeed, our data do not support a role for vitamin D in training-associated muscle plasticity and functionality, at least not in older adults (with and without moderate COPD) with suboptimal to adequate baseline levels of 25(OH)D. More precisely, vitamin D_3_ supplementation had no effect on core outcome domains such as changes in muscle strength, muscle mass, endurance performance and general muscle cell characteristics, and its effects on muscle transcriptome were largely limited to gene sets relating to endothelial and cardiovascular functions. The validity of this insight is fortified by our thorough methodological and analytical approach. This included accounting for previous methodological issues such as a lack of a pre-training supplementation period, low vitamin D dosages, neglecting to standardize test/training routines and characteristics such as supervision of training sessions, test-retest analyses of functional and biological outcome measures, familiarization to training and a low reproducibility of singular outcome measures. Our analytical approach also accounted for the potential confounding effects of the heterogeneity of the study population, as no interaction was found between effects of vitamin D_3_ supplementation and disease status (healthy vs COPD), and differences in pre-RCT vitamin D status, as all [25(OH)D]_baseline_ quartiles responded in similar manners.

Despite our substantial efforts to strengthen the ecological value of the data set, there are aspects of vitamin D biology that remain unresolved, and that may have affected the conclusions and outcomes of the study. For example, in skeletal muscle, adequate vitamin D signaling may occur at 25(OH)D levels lower than the defined cutoff (insufficient, < 50 nmol. L^-1^).^27^ Speculatively, all participants in the placebo arm may thus have been vitamin D-sufficient at the onset of resistance training, leaving our quartile-based analyses with limited biological value. Indeed, studies have suggested that vitamin D insufficiency will affect human muscle in an adverse manner only at concentrations < 30 nmol. L^-1^.^146^ One could also argue that the beneficial effects of vitamin D_3_ supplementation on muscular responses to resistance training demand a longer training period, or that the individual variations in responses to vitamin D supplementation and resistance training lowered the resolution of the data set, or that 25(OH)D is a poor proxy marker of vitamin D biology that does not reflect 1,25 dihydroxycholecalciferol levels, the metabolically active form of vitamin D.^147^ As for the latter, at present, total serum [25(OH)D] presents the standard measure to assess vitamin D status.^63^ However, it can be argued that the levels of free 25(OH)D (i.e. not bound to vitamin D binding protein or albumin; ∼0.03 %) represent a more accurate measure of vitamin D status in clinical settings.^148^ This assumption is supported by the notion that mice lacking vitamin D binding protein, and therefore display very low total 25(OH)D levels (∼8 mmol/L), do not show signs of vitamin D deficiency unless they were put on a vitamin D deficient diet.^149^ More research is clearly needed to elucidate on these perspectives. However, neither of these potential shortcomings can fully explain the near complete lack of vitamin D effects seen for training-associated changes in muscle biological characteristics, particularly those measured using RNA-seq. Indeed, in our transcriptome analyses, not a single gene was found to be vitamin D_3_-sensitive after a period of resistance training, which is surprising given the accepted dogma that vitamin D primarily acts as a transcriptional regulator.^56^ Moreover, gene sets that were identified as vitamin D_3_-sensitive in gene enrichment analyses were largely associated with vascular function rather than muscle cell biology.

Despite the general lack of effects of vitamin D_3_ supplementation on muscle mass and phenotype (primary objectives of the study), as well as the lack of effects on other muscle functional and biological traits, the data set contained a few interesting observations. First, the effects of vitamin D_3_ supplementation *per se* on expression of mitochondrial genes and the effects of combined vitamin D_3_ supplementation and resistance training on biomarkers of endothelial and vascular biology calls for further study. For example, these biological features would arguably be more decisive for adaptations to endurance-like training, posing the intriguing possibility that vitamin D_3_ supplementation may exert ergogenic effects on outcome of such training. Second, in participants with high baseline fat proportions/high BMI, vitamin D_3_ supplementation led to increased training-associated changes in muscle strength. In these participants, the bioavailability of vitamin D may thus have been compromised by the high fat content (in the placebo arm, though they did not exhibit lowered 25(OH)D levels), corroborating with previous observation of interactions between vitamin D biology and fat mass.^112^ While this may indicate that vitamin D exerts direct effects on muscle biology, as muscle strength is predominately defined by muscle mass,^150^ this seems unlikely as no such vitamin D_3_-effect was seen for other muscle-specific outcome measures (e.g. muscle mass and phenotype). The causality may thus involve other physiological adaptations such as motoneuron function,^151^ which has indeed been suggested to be affected by vitamin D supplementation in rodents.^152^ Third, vitamin D_3_ supplementation and resistance training led to differential levels of cortisol in blood compared to placebo. The biological significance of this is difficult to interpret, and it has not previously been observed by others. However, it may involve prevention of hypercalcemia,^136^ as previously discussed. Speculatively, the increase in cortisol may even have impaired any beneficial effects of vitamin D_3_ supplementation on muscle plasticity (perhaps together with calcium supplementation), as cortisol is known for its catabolic effects on a range of physiological systems.^17^ Once again, it seems timely to reiterate on the absence of vitamin D_3_ effects on muscle biology and transcriptome, as well as the high efficacy of the resistance training intervention on functional and biological characteristics in the data set as a whole, leaving impairing effects of cortisol/calcium on muscle characteristics as unlikely.

In conclusion, in old adults with or without COPD, vitamin D_3_ supplement efficiently improved vitamin D-status without any adverse effects, but did not lead to beneficial effects in resistance training-associated changes in muscle function or characteristics. This rejects the notion that vitamin D_3_ supplementation is necessary to obtain adequate muscular responses to resistance training in the general older population. Secondary analyses revealed positive effects of vitamin D_3_ supplementation for participants with high proportions of fat mass and in gene sets involved in vascular functions, thus advocating further research to elucidate on these specific biological characteristics. Finally, the training program was well-tolerated and associated with pronounced effects on a variety of health variables, emphasizing the use of resistance training for older subjects to relieve sarcopenia and to maintain functional capacity.

## Data Availability

All data generated or analysed during this study are included in this published article (and its supplementary information files).

## Acknowledgements

The authors would like to express their gratitude to students Katarina Alsvik, Karianne Pedersen, Nicolai Blindheim Jacobsen, Jan Christian Fjelltun, Håvard Berge Bredesen, Karoline Michalsen, Håkon Fure Jerpseth, Karin Heggeli Moen, Krister Flobergseter, Mads-Henrik Hafsmo, Sondre Bjerke, Joachim Skjeseth Fjeller, Vilde Bakkhaug, Vetle Olsen, Hilde Juvik, Hallvard Underdal, Anniken Braaten, Anette Gårderløkken, Malene Wilhelmsen, Vemund Lien, Malene Grahl-Madsen, Pauline Forren, Anders Kristoffersen, Jørn Klepp Thorgersen, Berit Hauge Aakvik, Ole-Jørgen Folkestad, Simen Bratberg Ramstad, Lasse Løwstrøm Aulin, Simen Næss Berge, Marius Midtmageli Bekkemellem, Martine Pedersen, Kristian Lian, Even Hovland Rosenlund, Henrik Eckhoff, Synne Skogstad, Marte Johannson, Simen Longva Hedlund, Jon Sindre Aas, Marte Fosvold Løtveit, Stine Studsrød and Marius Fagerås for invaluable assistance during intervention follow-up and data sampling. The authors are grateful for the contribution from Tore J. Rødølen (Granheim Lung Hospital, Innlandet Hospital Trust) and the technical support from Bente Malerbakken (Lillehammer Hospital for Rheumatic Diseases) and Randi Christiansen. We also acknowledge Pharma Nord ApS for a good cooperation and sponsoring the vitamin D_3_ and placebo supplements in the study. Further, the authors wish to thank Proteinfabrikken A/S for sponsoring the project with protein chocolate bars and Johanne Haugen, University of Bergen, for taking care of the third-party randomization. Last but not least, thanks to all participants for your cooperation. Without your effort and dedication, this study had not been possible.

## Online supporting material

Additional supporting information may be found online in the Supporting Information section.

Figure S1 General efficacy of the RCT

Figure S2 Sample-resample reliability measures of immunohistochemical assessments

Figure S3 Baseline vitamin D-status and the interaction with the study’s main outcomes

Figure S4 Baseline body fat proportions/body mass index and the interaction with the study’s main outcomes

Table S1 qPCR primer sequences and performance Table S2 Statistical summary table

Table S3 Statistical summary table, qPCR data

Table S4 Computed factors for main outcome domains

Table S5 Gene ontology analyses, effects of resistance training

Table S6 Gene ontology analyses, effects of combined vitamin D_3_ supplementation and resistance training Table S7 Gene ontology analyses, effects of vitamin D_3_ supplementation

Table S8 Blood and health variables

